# Impact of the COVID-19 pandemic on treatment patterns for US patients with metastatic solid cancer

**DOI:** 10.1101/2021.09.22.21263964

**Authors:** Ravi B. Parikh, Samuel U. Takvorian, Daniel Vader, E. Paul Wileyto, Amy S. Clark, Daniel J. Lee, Gaurav Goyal, Gabrielle B. Rocque, Efrat Dotan, Daniel M. Geynisman, Pooja Phull, Philippe E. Spiess, Roger Y. Kim, Amy J. Davidoff, Cary P. Gross, Natalia Neparidze, Rebecca A. Miksad, Gregory S. Calip, Caleb M. Hearn, Will Ferrell, Lawrence N. Shulman, Ronac Mamtani, Rebecca A. Hubbard, for the PRACTICE Investigators

## Abstract

**Background:** The COVID-19 pandemic has led to delays in patients seeking care for life-threatening conditions; however, its impact on treatment patterns for patients with metastatic cancer is unknown. We assessed the COVID-19 pandemic’s impact on time to treatment initiation (TTI) and treatment selection for patients newly diagnosed with metastatic solid cancer.

**Methods:** We used an electronic health record-derived longitudinal database curated via technology-enabled abstraction to identify 14,136 US patients newly diagnosed with de novo or recurrent metastatic solid cancer between January 1 and July 31 in 2019 or 2020. Patients received care at ∼280 predominantly community-based oncology practices. Controlled interrupted time series analyses assessed the impact of the COVID-19 pandemic period (April-July 2020) on TTI, defined as the number of days from metastatic diagnosis to receipt of first-line systemic therapy, and use of myelosuppressive therapy.

**Results:** The adjusted probability of treatment within 30 days of diagnosis [95% confidence interval] was similar across periods: January-March 2019 41.7% [32.2%, 51.1%]; April-July 2019 42.6% [32.4%, 52.7%]; January-March 2020 44.5% [30.4%, 58.6%]; April-July 2020 46.8% [34.6%, 59.0%]; adjusted percentage-point difference-in-differences 1.4% [-2.7%, 5.5%]. Among 5,962 patients who received first-line systemic therapy, there was no association between the pandemic period and use of myelosuppressive therapy (adjusted percentage-point difference-in-differences 1.6% [-2.6%, 5.8%]). There was no meaningful effect modification by cancer type, race, or age.

**Conclusions:** Despite known pandemic-related delays in surveillance and diagnosis, the COVID-19 pandemic did not impact time to treatment initiation or treatment selection for patients with metastatic solid cancers.

## INTRODUCTION

The COVID-19 pandemic has led to declines in patients seeking care for life-threatening conditions, such as acute myocardial infarction and stroke, as well as care delays for screening and management of chronic medical conditions.^1–5^ For patients with cancer, who may be particularly vulnerable to COVID-19 infection,^6–8^ early research suggested changes in practice patterns leading to care delays and treatment modifications.^9–17^ Some of these changes were supported by guidelines issued during the pandemic,^18^ which encouraged consideration of non-myelosuppressive regimens despite mixed evidence linking the risk and severity of COVID-19 infection to immunosuppression from cancer therapy.^8,19–21^ These care disruptions may have been particularly prominent for patients with metastatic cancer, for whom treatments are palliative rather than curative. A recent systematic review identified 62 studies evaluating pandemic-related delays across the cancer care continuum; however, the majority of these studies used single-institution data and did not focus on patients with metastatic cancer.^22^ Thus, little is known about the impact of the pandemic on changes in treatment patterns for patients with metastatic cancer.

Because treatment delays cause patient distress and are associated with increased mortality for patients with cancer,^23–27^ time to treatment initiation (TTI) is a patient-centered quality metric and outcome that has been used to evaluate the impact of health policies on cancer care.^9,28,29^ TTI may also serve as a barometer of capacity limitation and care delivery disruption during the COVID-19 pandemic.^30–34^ Moreover, pandemic-related delays or changes in cancer treatment may have disproportionately affected minority groups including African-American patients, who even prior to the pandemic were less likely to receive guideline-concordant systemic therapy for metastatic cancer than White patients.^35–40^ It is thus critical to identify whether the COVID-19 pandemic resulted in changes in treatment patterns for patients with metastatic cancer, with potential downstream consequences that could adversely affect patient outcomes and equitable cancer care.

The objective of this study was to evaluate the impact of the COVID-19 pandemic on time to treatment initiation and treatment selection for patients newly diagnosed with metastatic solid cancer, with attention to race- and age-based disparities. We hypothesized that the pandemic would be associated with delays in initiation of systemic therapy and increased use of non-myelosuppressive therapies.

## METHODS

### Study design

We applied a retrospective controlled interrupted time series approach to evaluate associations between the COVID-19 pandemic period and changes in TTI and use of myelosuppressive therapy. The study adhered to Strengthening the Reporting of Observational Studies in Epidemiology (STROBE) reporting guidelines and was exempted by the University of Pennsylvania and WCG Institutional Review Boards prior to study conduct owing to use of de-identified data only.

### Data source

This study used the nationwide Flatiron Health database, an electronic health record (EHR)-derived, longitudinal database comprising de-identified patient-level structured and unstructured data, curated via technology-enabled abstraction.^41,42^ During the study period, data originated from approximately 280 US cancer clinics (∼800 sites of care). The majority of patients in the database originated from community oncology settings. The data were de-identified and subject to obligations to prevent re-identification and protect patient confidentiality.

### Participants

The main study sample included adult patients (age ≥ 18 years) with a new diagnosis of metastatic solid cancer from January 1-July 31, 2019 or January 1-July 31, 2020. Metastatic status was determined using both structured data and abstracted unstructured data from clinical, imaging and pathology notes, and included de novo (M1 at initial diagnosis) or recurrent (M0 at initial diagnosis) diagnoses. Eligible cancer types were breast, colorectal, non-small cell lung (NSCLC), pancreas, prostate, renal cell, or urothelial cancer. We excluded patients with incomplete historical treatment data (defined as 90 days or more between diagnosis and the earliest structured activity documented in the EHR [n=1,631]), fewer than two documented clinical visits after metastatic cancer diagnosis (n=1,275), multiple metastatic malignancies (n=66), first-line treatment starting prior to recorded metastatic diagnosis date (n=682), or receiving therapy that was not part of National Comprehensive Cancer Network guidelines (n=344). We also excluded patients diagnosed during a 30-day “washout” period (March 8 to April 7) encompassing the start of most state stay-at-home orders in 2020 (**eTable 1**), and historical controls during the comparable period in 2019 (n=2,127). **eFigure 1** illustrates our cohort selection.

We evaluated changes in treatment selection in a sub-sample of patients diagnosed with metastatic breast, NSCLC, prostate, or urothelial cancer during the study period who received a systemic therapy within 60 days of metastatic diagnosis (n=6,721). We selected these four cancers because they have guideline-based myelosuppressive and non-myelosuppressive options for frontline therapy. Furthermore, frontline treatment guidelines^43–46^ for these metastatic cancers did not change substantially during the study period, allowing for comparisons to historical controls. In addition to exclusions applied to the main study sample, patients were excluded if they received first-line treatment directed at a targetable mutation (EGFR, ALK, ROS-1, or BRAF for NSCLC; HER-2 for breast) or microsatellite instability (n=759). These patients were excluded because their treatment decisions were likely influenced by the presence of an actionable genetic or molecular aberration rather than by factors related to the pandemic.

### Main outcomes and measures

The primary outcome was time to treatment initiation (TTI), defined as the number of days from metastatic diagnosis to receipt of first-line systemic therapy. Patients were censored at their last structured activity within the Flatiron Health network (defined as the latest date of a clinical visit, laboratory check, or treatment receipt) or 90 days after diagnosis, whichever occurred first. The secondary outcome was receipt of myelosuppressive treatment. Myelosuppressive treatment was defined as any regimen containing cytotoxic chemotherapy or a cyclin-dependent kinase (CDK) inhibitor. Checkpoint inhibitors (NSCLC, urothelial) and hormone therapies (breast, prostate) without concurrent myelosuppressive therapy were considered non-myelosuppressive (see **eTable 2** for treatment categorizations).

The primary exposure was time period (April 8-July 31 vs. January 1-March 8) and year (2020 vs. 2019) of metastatic cancer diagnosis. These intervals corresponded with time periods in 2020 when the COVID-19 pandemic would be more vs. less likely to influence patient treatment based on the date of most states’ stay-at-home orders. In our controlled interrupted time series approach, the comparison of interest was defined as the change in TTI (or receipt of myelosuppressive therapy) across time periods in 2020 compared to the change across time periods in 2019.

Covariates included age, gender, race (non-Hispanic White, non-Hispanic Black, Hispanic, or other), insurance type (commercial, government, or other), Eastern Cooperative Oncology Group (ECOG) performance status (<2 or ≥2), documented opioid medication order (yes or no), calendar day of metastatic cancer diagnosis, and cancer type. All covariates were ascertained at the time of metastatic cancer diagnosis.

Missing baseline covariate data were accounted for using multiple imputation via chained equations with 10 imputations. Continuous variables were imputed using an approach that allowed for heterogeneous within-group variance by practice.^47^ Categorical and dichotomous variables were imputed using multinomial logistic regression and logistic regression, respectively.

### Statistical methods

Frequencies and proportions of baseline characteristics were summarized by time period. Standardized mean differences were used to describe differences in baseline characteristics across the 4 time periods; a standardized mean difference >0.1 was considered a meaningful difference.^48^ The Kaplan-Meier estimator was used to estimate unadjusted median TTI within each time period. We conducted adjusted analyses of TTI using Cox proportional hazards regression. The primary exposure was an interaction between period (April-July vs. January-March) and year (2020 vs. 2019) of metastatic cancer diagnosis. All models were adjusted for age, sex, race, insurance, ECOG, opioid prescription, a linear time trend for calendar day of metastatic cancer diagnosis, and cancer type, and used robust standard errors to allow for within-practice correlation. Our primary analysis included an additional three-way interaction between period, year, and cancer type, to investigate effect modification by cancer type. Exploratory analyses excluded the cancer type interaction and included three-way interactions between period, year, and race or, in a separate model, period, year, and age group, to investigate effect modification by race or age group, respectively. After fitting the Cox models, we used marginal standardization to estimate the predicted probabilities of treatment within 30 days of metastatic cancer diagnosis within each time-period. Estimates across the 10 imputations were combined using Rubin’s rules.^47,49^

Analyses of the sub-sample of patients who initiated treatment within 60 days of diagnosis employed a similar approach using logistic regression rather than Cox regression to model use of myelosuppressive therapy (vs not). Marginal standardization was applied to logistic regression estimates to obtain adjusted probabilities of receiving myelosuppressive therapy.

### Sensitivity analyses

We performed a sensitivity analysis to verify the robustness of our findings to an alternate definition of pandemic period exposure. Rather than defining one exposure period for all study participants that encompassed most state stay-at-home orders, we varied the exposure period for individual participants based on the stay-at-home order date of their state of residence (see **eTable 1** for dates), thereby defining variable 30-day washout periods by state.

Data analyses were conducted between November 2020 and April 2021 using R, version 4.0.4. All hypothesis tests were two-tailed with alpha equal to 0.05. Missing data were imputed using the mice package, version 3.13.0.^50^ Cox Proportional hazards models were fit using the survival package, version 3.2.11^51^ and regression standardization conducted using stdReg, version 3.4.1^52^. All analytic code is available at https://github.com/PRACTICE-research-group/COVID19-treatment-patterns.

## RESULTS

### Baseline characteristics

**Table 1** shows the distribution of patient characteristics in the main study sample by time period and year. Of 14,136 patients with documented newly diagnosed metastatic solid cancer during the study period, 2,954 (20.9%) were diagnosed from January-March 2019, 4,745 (33.6%) from April-July 2019, 2,640 (18.7%) from January-March 2020, and 3,797 (26.9%) from April-July 2020. There were no meaningful differences in the distributions of age, gender, race, insurance, practice setting, and performance status by time period within each year (standardized mean differences [SMDs] <0.1). The most common cancers were NSCLC (41.3%), colorectal (18.4%), and breast (11.6%) cancers; there were no differences in the distribution of cancers by time period. Overall, 62.9% of patients were diagnosed with de novo metastatic disease; as a proportion of overall new metastatic cancer diagnoses, de novo metastatic diagnoses were more common in the COVID-19 period (April-July 2020 67.0%) than in the pre-COVID-19 periods (January-March 2019 61.2%; April-July 2019 61.5%; January-March 2020 61.5%; SMD 0.11). **eTable 3** describes the sub-sample of patients (n=5,962) who were diagnosed with metastatic NSCLC, breast, prostate, or urothelial cancer and treated within 60 days of diagnosis. The distribution of baseline characteristics in this sub-sample was similar to the full cohort.

**Table 1.** Population characteristics

|  | 2019 |  |  |  |  | 2020 |  |  |  |  | Total |  |
| --- | --- | --- | --- | --- | --- | --- | --- | --- | --- | --- | --- | --- |
|  | Jan 1-Mar 8 |  | Apr 8-Jul 31 |  | SMD | Jan 1-Mar 8 |  | Apr 8-Jul 31 |  | SMD |  |  |
| n | 2954 |  | 4745 |  |  | 2640 |  | 3797 |  |  | 14136 |  |
| Cancer (n, %) |  |  |  |  |  |  |  |  |  |  |  |  |
| Breast | 382 | (12.9%) | 589 | (12.4%) | 0.089 | 282 | (10.7%) | 393 | (10.4%) | 0.066 | 1646 | (11.6%) |
| Colorectal | 553 | (18.7%) | 862 | (18.2%) |  | 466 | (17.7%) | 722 | (19.0%) |  | 2603 | (18.4%) |
| NSCLC | 1170 | (39.6%) | 1987 | (41.9%) |  | 1122 | (42.5%) | 1562 | (41.1%) |  | 5841 | (41.3%) |
| Pancreatic | 261 | (8.8%) | 388 | (8.2%) |  | 251 | (9.5%) | 368 | (9.7%) |  | 1268 | (9.0%) |
| Prostate | 282 | (9.5%) | 383 | (8.1%) |  | 214 | (8.1%) | 347 | (9.1%) |  | 1226 | (8.7%) |
| RCC | 133 | (4.5%) | 274 | (5.8%) |  | 147 | (5.6%) | 178 | (4.7%) |  | 732 | (5.2%) |
| UCC | 173 | (5.9%) | 262 | (5.5%) |  | 158 | (6.0%) | 227 | (6.0%) |  | 820 | (5.8%) |
| Median Age (IQR) | 70 | (61, 77) | 70 | (61, 77) | 0.013 | 69 | (61, 77) | 69 | (62, 77) | 0.008 | 70 | (61, 77) |
| Gender (n, %) |  |  |  |  |  |  |  |  |  |  |  |  |
| Female | 1330 | (45.0%) | 2295 | (48.4%) | 0.067 | 1250 | (47.3%) | 1784 | (47.0%) | 0.007 | 6659 | (47.1%) |
| Male | 1624 | (55.0%) | 2450 | (51.6%) |  | 1390 | (52.7%) | 2013 | (53.0%) |  | 7477 | (52.9%) |
| Race (n, %) |  |  |  |  |  |  |  |  |  |  |  |  |
| Non-Hispanic White | 1801 | (68.7%) | 2912 | (69.1%) | 0.031 | 1554 | (66.8%) | 2211 | (66.3%) | 0.015 | 8478 | (67.8%) |
| Non-Hispanic Black | 270 | (10.3%) | 458 | (10.9%) |  | 242 | (10.4%) | 355 | (10.6%) |  | 1325 | (10.6%) |
| Hispanic | 164 | (6.3%) | 240 | (5.7%) |  | 131 | (5.6%) | 197 | (5.9%) |  | 732 | (5.9%) |
| Other | 386 | (14.7%) | 606 | (14.4%) |  | 400 | (17.2%) | 571 | (17.1%) |  | 1963 | (15.7%) |
| Missing | 333 |  | 529 |  |  | 313 |  | 463 |  |  | 1638 |  |
| Insurance (n, %) |  |  |  |  |  |  |  |  |  |  |  |  |
| Commercial | 1435 | (48.6%) | 2315 | (48.8%) | 0.048 | 1343 | (50.9%) | 1915 | (50.4%) | 0.026 | 7008 | (49.6%) |
| Government | 585 | (19.8%) | 1015 | (21.4%) |  | 556 | (21.1%) | 775 | (20.4%) |  | 2931 | (20.7%) |
| Unknown/not documented/self-pay | 934 | (31.6%) | 1415 | (29.8%) |  | 741 | (28.1%) | 1107 | (29.2%) |  | 4197 | (29.7%) |
| Practice type (n, %) |  |  |  |  |  |  |  |  |  |  |  |  |
| Academic | 279 | (9.4%) | 471 | (9.9%) | 0.016 | 235 | (8.9%) | 341 | (9.0%) | 0.003 | 1326 | (9.4%) |
| Community | 2675 | (90.6%) | 4274 | (90.1%) |  | 2405 | (91.1%) | 3456 | (91.0%) |  | 12810 | (90.6%) |
| ECOG performance status (n, %) |  |  |  |  |  |  |  |  |  |  |  |  |
| 0-1 | 1176 | (82.1%) | 1816 | (82.1%) | 0.001 | 1086 | (83.5%) | 1540 | (81.7%) | 0.047 | 5618 | (82.3%) |
| >=2 | 257 | (17.9%) | 396 | (17.9%) |  | 214 | (16.5%) | 344 | (18.3%) | 0.047 | 1211 | (17.7%) |
| Missing | 1521 |  | 2533 |  |  | 1340 |  | 1913 |  |  | 7307 |  |
| Opioid prescription (n, %) |  |  |  |  |  |  |  |  |  |  |  |  |
|  | 220 | (7.4%) | 362 | (7.6%) | 0.007 | 179 | (6.8%) | 274 | (7.2%) | 0.017 | 1035 | (7.3%) |
| De novo metastatic (n, %) |  |  |  |  |  |  |  |  |  |  |  |  |
|  | 1647 | (61.2%) | 2676 | (61.5%) | 0.006 | 1491 | (61.5%) | 2329 | (67.0%) | 0.113 | 8143 | (62.9%) |
| Missing | 261 |  | 391 |  |  | 217 |  | 319 |  |  | 1188 |  |

NOTE: SMD = Standardized mean difference; NSCLC = Non-small cell lung carcinoma; RCC = renal cell carcinoma; UCC = urothelial cell carcinoma; ECOG = Eastern Cooperative Oncology Group

### Time to treatment initiation

Across all periods, the median time to systemic treatment initiation was 35 days with 44.0% (95% confidence interval [CI] 43.2%, 44.8%) of patients initiating treatment within 30 days of metastatic diagnosis. Unadjusted and adjusted probabilities of treatment initiation within 30 days are shown in **Table 2**. In our primary analysis, the difference in the proportion of patients initiating treatment within 30 days in April-July compared to January-March was similar in 2019 and 2020 (adjusted probability of treatment within 30 days [95% CI]: January-March 2019 41.7% [32.2%, 51.1%]; April-July 2019 42.6% [32.4%, 52.7%]; January-March 2020 44.5% [30.4%, 58.6%]; April-July 2020 46.8% [34.6%, 59.0%]; adjusted percentage-point difference-in-differences 1.4% [-2.7%, 5.5%]) (**Table 2**). There was no evidence of effect modification by cancer type (interaction p=0.247) (**Figure 1**), race (**eTable 4**; p=0.100), or age (**eTable 5**; p=0.653).

**Table 2.**
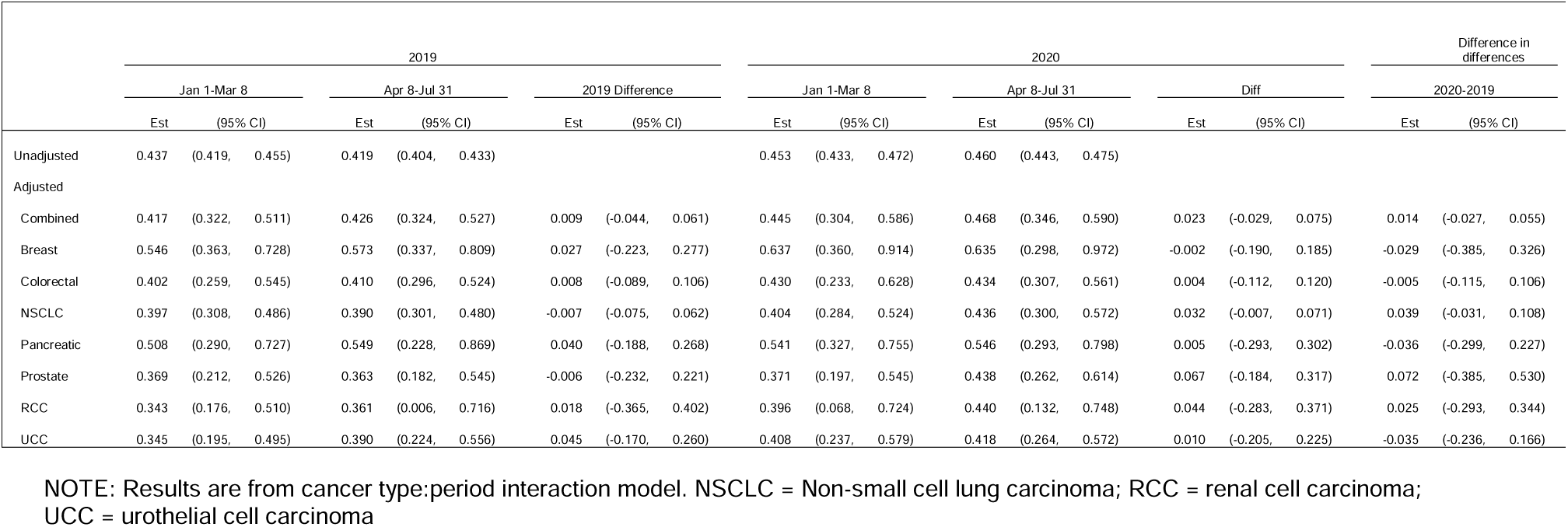
Adjusted probability of treatment within 30 days

|  | 2019 |  |  |  |  |  | 2020 |  |  |  |  |  | Difference in differences |  |
| --- | --- | --- | --- | --- | --- | --- | --- | --- | --- | --- | --- | --- | --- | --- |
|  | Jan 1-Mar 8 |  | Apr 8-Jul 31 |  | 2019 Difference |  | Jan 1-Mar 8 |  | Apr 8-Jul 31 |  | Diff |  | 2020-2019 |  |
|  | Est | (95% CI) | Est | (95% CI) | Est | (95% CI) | Est | (95% CI) | Est | (95% CI) | Est | (95% CI) | Est | (95% CI) |
| Unadjusted | 0.437 | (0.419, 0.455) | 0.419 | (0.404, 0.433) |  |  | 0.453 | (0.433, 0.472) | 0.460 | (0.443, 0.475) |  |  |  |  |
| Adjusted |  |  |  |  |  |  |  |  |  |  |  |  |  |  |
| Combined | 0.417 | (0.322, 0.511) | 0.426 | (0.324, 0.527) | 0.009 | (-0.044, 0.061) | 0.445 | (0.304, 0.586) | 0.468 | (0.346, 0.590) | 0.023 | (-0.029, 0.075) | 0.014 | (-0.027, 0.055) |
| Breast | 0.546 | (0.363, 0.728) | 0.573 | (0.337, 0.809) | 0.027 | (-0.223, 0.277) | 0.637 | (0.360, 0.914) | 0.635 | (0.298, 0.972) | -0.002 | (-0.190, 0.185) | -0.029 | (-0.385, 0.326) |
| Colorectal | 0.402 | (0.259, 0.545) | 0.410 | (0.296, 0.524) | 0.008 | (-0.089, 0.106) | 0.430 | (0.233, 0.628) | 0.434 | (0.307, 0.561) | 0.004 | (-0.112, 0.120) | -0.005 | (-0.115, 0.106) |
| NSCLC | 0.397 | (0.308, 0.486) | 0.390 | (0.301, 0.480) | -0.007 | (-0.075, 0.062) | 0.404 | (0.284, 0.524) | 0.436 | (0.300, 0.572) | 0.032 | (-0.007, 0.071) | 0.039 | (-0.031, 0.108) |
| Pancreatic | 0.508 | (0.290, 0.727) | 0.549 | (0.228, 0.869) | 0.040 | (-0.188, 0.268) | 0.541 | (0.327, 0.755) | 0.546 | (0.293, 0.798) | 0.005 | (-0.293, 0.302) | -0.036 | (-0.299, 0.227) |
| Prostate | 0.369 | (0.212, 0.526) | 0.363 | (0.182, 0.545) | -0.006 | (-0.232, 0.221) | 0.371 | (0.197, 0.545) | 0.438 | (0.262, 0.614) | 0.067 | (-0.184, 0.317) | 0.072 | (-0.385, 0.530) |
| RCC | 0.343 | (0.176, 0.510) | 0.361 | (0.006, 0.716) | 0.018 | (-0.365, 0.402) | 0.396 | (0.068, 0.724) | 0.440 | (0.132, 0.748) | 0.044 | (-0.283, 0.371) | 0.025 | (-0.293, 0.344) |
| UCC | 0.345 | (0.195, 0.495) | 0.390 | (0.224, 0.556) | 0.045 | (-0.170, 0.260) | 0.408 | (0.237, 0.579) | 0.418 | (0.264, 0.572) | 0.010 | (-0.205, 0.225) | -0.035 | (-0.236, 0.166) |

NOTE: Results are from cancer type:period interaction model. NSCLC = Non-small cell lung carcinoma; RCC = renal cell carcinoma; UCC = urothelial cell carcinoma

**Figure 1.**
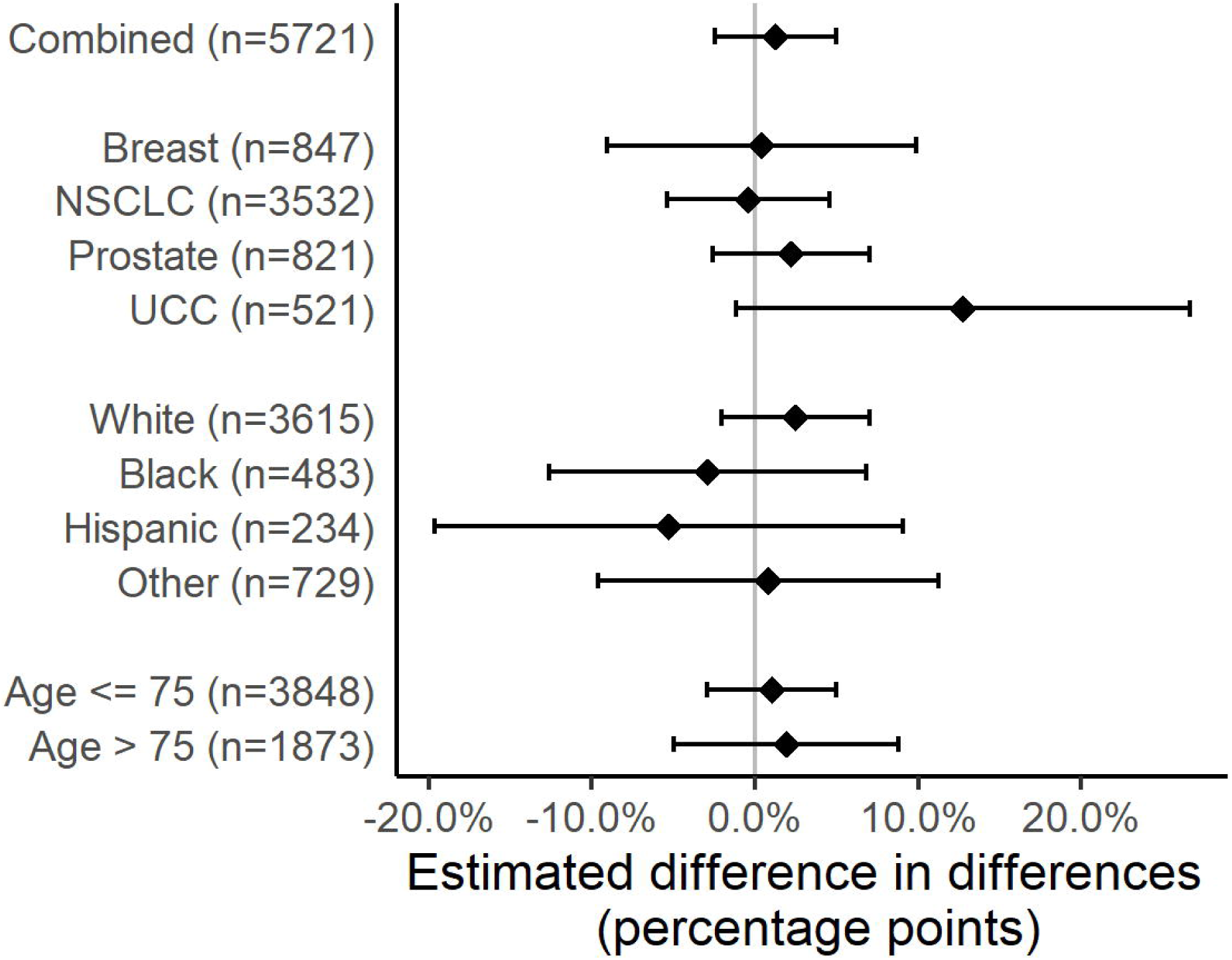
Changes in the adjusted probability of treatment initiation within 30 days of metastatic diagnosis between COVID-19 and pre-COVID-19 periods. Figure 1 displays the differential effect of the COVID-19 period on the probability of 30-day treatment initiation by cancer type, race, and age among patients with newly diagnosed de novo or recurrent metastatic solid cancer.

### Treatment selection

Among the 5,962 patients who received first-line systemic therapy within 60 days of diagnosis, 67.2% received myelosuppressive therapy (range 3.2% [prostate cancer] to 81.0% [breast cancer]). The difference in the adjusted probability of receiving myelosuppressive therapy in April-July compared to January-March was similar in 2019 and 2020 (January-March 2019 69.8% [65.1%, 74.4%]; April-July 2019 66.7% [60.9%, 72.5%]; January-March 2020 68.3% [65.1%, 71.4%]; April-July 66.8% [63.3%, 70.2%]; adjusted percentage-point difference-in-differences 1.6% [-2.6%, 5.8%]) (**Table 3**). There was no evidence of effect modification by cancer type (p=0.209) (**Figure 2**), race (**eTable 6**; p=0.130), or age (**eTable 7**; p=0.483).

**Table 3.**
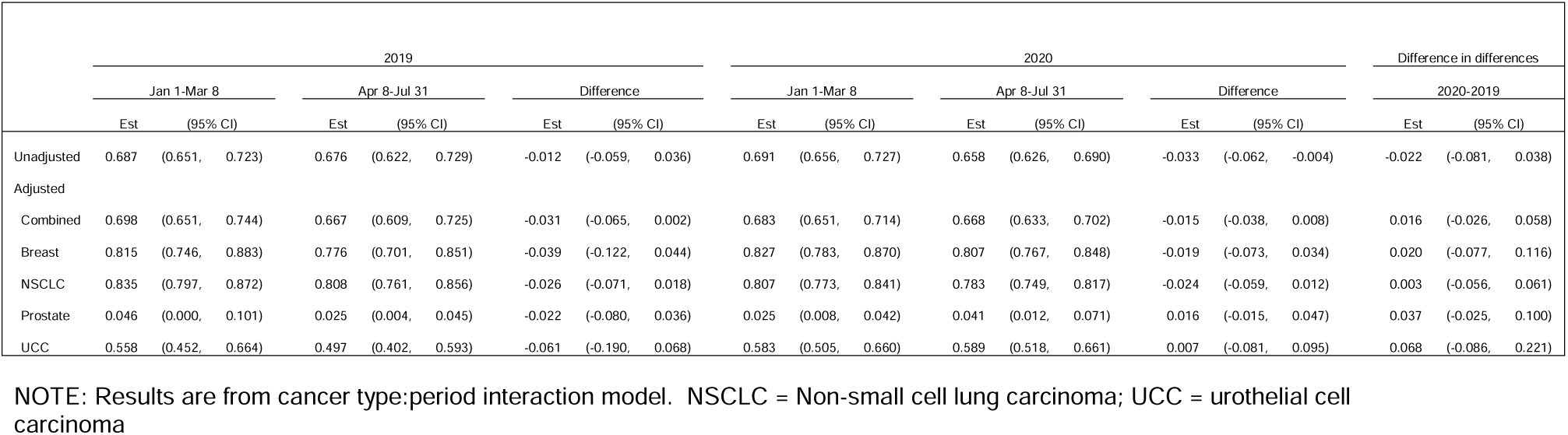
Adjusted probabilities of receipt of myelosuppressive therapy

**Figure 2.**
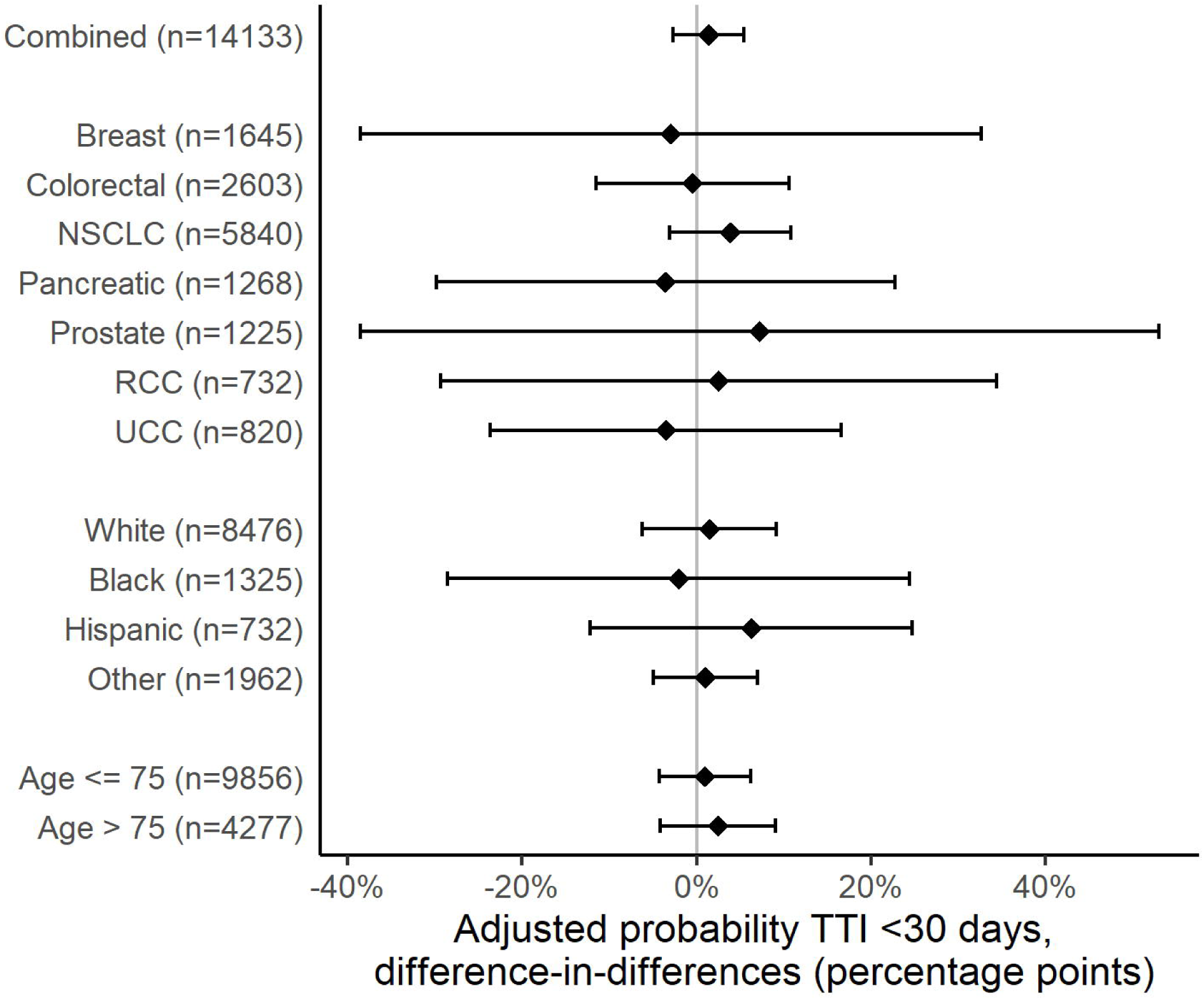
Changes in the adjusted probability of receiving myelosuppressive therapy after metastatic diagnosis between COVID-19 and pre-COVID-19 periods. Figure 2 displays the differential effect of the COVID-19 period on the probability of receiving myelosuppressive therapy by cancer type, race, and age among patients with newly diagnosed de novo or recurrent metastatic solid cancer.

### Sensitivity analyses

Results from a sensitivity analysis using a state-specific exposure definition based on dates of state stay-at-home orders were consistent with results from the primary analysis (**eTables 8-9**).

## DISCUSSION

In this large, multi-site cohort of patients with metastatic solid cancer, we assessed the impact of the COVID-19 pandemic on time to treatment initiation and treatment selection using a quasi-experimental approach. We did not find evidence that the pandemic period was associated with delayed systemic therapy or increased use of non-myelosuppressive therapy. We did observe changes in disease presentation during the COVID-19 period – most notably, an increased proportion of patients presenting with de novo metastatic disease. Our analysis suggests that previously reported pandemic-associated diagnostic delays may have resulted in more acute presentations of metastatic disease but not delays in systemic treatment initiation or preference against use of myelosuppressive therapies.

Our findings stand in contrast to earlier studies evaluating COVID-19 pandemic-related disruptions in cancer care, which found evidence of care delays across the cancer continuum.^11,13,17^ Several factors may account for this discrepancy. First, previously reported declines in cancer screening and diagnoses may have contributed to greater available capacity in outpatient clinics and infusion suites for those needing prompt treatment.^16^ Second, we observed a 5-6 percentage-point increase in the proportion of de novo metastatic diagnoses in the COVID-19 period compared to pre-COVID periods. Relative to recurrent metastatic diagnoses, which are often detected via routine surveillance imaging or laboratory testing when patients may not be symptomatic, de novo metastatic diagnoses are associated with greater symptomatic burden and worse overall mortality.^53^ It is possible that known pandemic-related decreases in routine imaging and laboratory surveillance contributed to the observed relative increase in presentation of potentially more symptomatic de novo metastatic diagnoses, which has been suggested in prior single-institution studies.^51^ Consequently, any pandemic-related delays in treatment initiation may have been balanced by the need for quicker treatment initiation for more symptomatic cases. Future studies with longer follow-up will be necessary to evaluate whether the relative increase of de novo presentations will persist, and what the consequences of this potential shift will be on future cancer-related outcomes. Nevertheless, delays in detection and diagnosis of recurrent metastatic disease during the early phase of the COVID-19 pandemic may be a harbinger for increased rates of symptomatic metastatic disease and cancer-associated mortality in later stages of the pandemic.

We did not find evidence of changes in the type of treatment selected, despite early professional society guidance in some cases cautioning against use of myelosuppressive therapy.^18^ The mechanisms behind this finding are unclear. An increased proportion of de novo metastatic diagnoses presenting with symptomatic disease may have led more physicians and patients than expected to prefer chemotherapy to achieve rapid debulking and disease control.^54^ Additionally, evidence emerged during the pandemic suggesting that myelosuppressive therapies might not, as initially suspected, be associated with increased COVID-19 severity or mortality among patients with cancer.^8^ Oncologists may have thus grown more comfortable with using myelosuppressive therapy during the pandemic period.

Our study has several advantages compared to prior studies examining pandemic-related treatment delays. First, we studied a large national cohort using electronic health record-derived data with minimal data lag, allowing for broad geographic coverage that accounted for state-specific stay-at-home orders, strong representation of community oncology practices, and greater data recency compared to other administrative databases. Second, we used a real-world dataset that harnesses technology-enabled chart abstraction to ascertain diagnoses and treatments, rather than relying solely on administrative claims from the COVID-19 pandemic period, which may be subject to measurement error and data lag.^55,56^ Finally, we used a quasi-experimental design to account for temporal confounding, such as known seasonal patterns in diagnoses and treatment-seeking behavior.^57^

### Limitations

Our study has several limitations. First, it is a retrospective study of a sample of predominantly community-based US oncology practices, and therefore our findings may not be reflective of all oncology practice. However, this database has been shown to be broadly representative of US oncology practices and patients.^41^ Second, outpatient EHR data may incompletely capture important variables that contribute to treatment patterns, such as patient preference or comorbidities, thus raising the possibility of unmeasured confounding. However, our quasi-experimental approach should account for these unmeasured confounders, assuming such confounders were consistent across time periods. Third, while we used the most up-to-date data available, there may be COVID-related delays in data capture affecting completeness of data from more recent time-periods. In particular, the pandemic could impact capture of metastatic cancer diagnoses. While this remains a hypothetical concern, future analyses should address this possibility. Fourth, our cohort was limited by a relatively small proportion of racial minorities and those with noncommercial insurance. This may have resulted in limited power for analyses of race- or age-based interactions, though notably there was some, non-statistically significant, evidence of delayed treatment among African-American patients. Given the disproportionate impact of the pandemic on care for minority groups, future analyses with larger, more diverse cohorts are needed. Finally, while we did not find any delays in systemic therapy initiation, our study did not address possible changes in systemic therapy dosing or schedules that may have occurred.

## Conclusions

In this large, nationwide study of patients newly diagnosed with metastatic solid cancer, we did not find evidence of treatment delays or preferential use of non-myelosuppressive therapies associated with the COVID-19 pandemic. An increased proportion of patients presenting with de novo metastatic cancers during the pandemic may portend a backlog of recurrent metastatic diagnoses stemming from pandemic-related delays in surveillance and diagnosis. Future studies with longer follow-up should assess whether COVID-related delays in presentation affect cancer-related outcomes among patients with metastatic cancers.

## Supporting information

PRACTICE Supplemental File

## Data Availability

All de-identified data generated or analyzed during this study is available upon request.

## NOTES

### Author disclosures

Ravi Parikh and Ronac Mamtani have received reimbursement from Flatiron, Inc. for travel and speaking. Rebecca Miksad and Gregory Calip report employment at Flatiron Health, Inc. There are no other conflicts of interest relevant to this submitted work.

### Author contributions

All authors have directly participated in the planning, execution, or analysis of the study, and have approved the final version of this manuscript.

### Disclaimers

The views expressed in this article are those of the authors, and no official endorsement by the National Cancer Institute, National Institutes for Health, or the Department of Health and Human Services is intended or should be inferred.

### Access to data and data analysis

Dr. Parikh, Dr. Takvorian, and Dr. Vader had full access to all the data in the study and take responsibility for the integrity of the data and the accuracy of the data analysis.

### Prior presentations

This was previously presented as a Poster Presentation at the 2021 ASCO Annual Meeting (Virtual).

### Funding

This study was supported by the National Cancer Institute K08-CA-263541-01 (to RBP)

### Role of the funder

The funders had no role in the design of the study; the collection, analysis, and interpretation of the data; the writing of the manuscript; and the decision to submit the manuscript for publication.

### Data availability

All de-identified data generated or analyzed during this study is available upon request.

## Acknowledgements

NA

## Notes

### Author Declarations

The University of Pennsylvania's Perelman School of Medicine IRB approved this study

## REFERENCES

1. Baum A, Schwartz MD. Admissions to Veterans Affairs Hospitals for Emergency Conditions During the COVID-19 Pandemic. JAMA. 2020;324(1):96. doi:10.1001/jama.2020.9972

2. Kansagra AP, Goyal MS, Hamilton S, Albers GW. Collateral Effect of Covid-19 on Stroke Evaluation in the United States. N Engl J Med. 2020;383(4):400–401. doi:10.1056/NEJMc2014816

3. Czeisler ME, Marynak K, Clarke KEN, et al. Delay or Avoidance of Medical Care Because of COVID-19-Related Concerns--United States, June 2020. Morbidity and Mortality Weekly Report. September 11, 2020:1250+.

4. Garcia S, Albaghdadi MS, Meraj PM, et al. Reduction in ST-Segment Elevation Cardiac Catheterization Laboratory Activations in the United States During COVID-19 Pandemic. J Am Coll Cardiol. 2020;75(22):2871–2872. doi:10.1016/j.jacc.2020.04.011

5. Solomon MD, McNulty EJ, Rana JS, et al. The Covid-19 Pandemic and the Incidence of Acute Myocardial Infarction. N Engl J Med. 2020;383(7):691–693. doi:10.1056/NEJMc2015630

6. Gupta S, Hayek SS, Wang W, et al. Factors Associated With Death in Critically Ill Patients With Coronavirus Disease 2019 in the US. JAMA Intern Med. 2020;180(11):1436–1446. doi:10.1001/jamainternmed.2020.3596

7. Kuderer NM, Choueiri TK, Shah DP, et al. Clinical impact of COVID-19 on patients with cancer (CCC19): a cohort study. The Lancet. 2020;395(10241):1907–1918. doi:10.1016/S0140-6736(20)31187-9

8. Lee LY, Cazier J-B, Angelis V, et al. COVID-19 mortality in patients with cancer on chemotherapy or other anticancer treatments: a prospective cohort study. Lancet Lond Engl. 2020;395(10241):1919–1926. doi:10.1016/S0140-6736(20)31173-9

9. Hawrot K, Shulman LN, Bleiweiss IJ, et al. Time to Treatment Initiation for Breast Cancer During the 2020 COVID-19 Pandemic. JCO Oncol Pract. 2021;(Journal Article):OP2000807-OP2000807. doi:10.1200/OP.20.00807

10. Perkons N, Kim C, Boedec C, et al. Quantifying the impact of the COVID-19 pandemic on gastrointestinal cancer care delivery. J Clin Oncol. 2021;39(3_suppl):30-30. doi:10.1200/JCO.2021.39.3_suppl.30

11. Papautsky EL, Hamlish T. Patient-reported treatment delays in breast cancer care during the COVID-19 pandemic. Breast Cancer Res Treat. 2020;184(1):249–254. doi:10.1007/s10549-020-05828-7

12. Schrag D, Hershman DL, Basch E. Oncology Practice During the COVID-19 Pandemic. JAMA. 2020;323(20):2005–2006. doi:10.1001/jama.2020.6236

13. Patt D, Gordan L, Diaz M, et al. Impact of COVID-19 on Cancer Care: How the Pandemic Is Delaying Cancer Diagnosis and Treatment for American Seniors. JCO Clin Cancer Inform. 2020;4:1059–1071. doi:10.1200/CCI.20.00134

14. Elkrief A, Kazandjian S, Bouganim N. Changes in Lung Cancer Treatment as a Result of the Coronavirus Disease 2019 Pandemic. JAMA Oncol. 2020;6(11):1805–1806. doi:10.1001/jamaoncol.2020.4408

15. Satish T, Raghunathan R, Prigoff JG, et al. Care Delivery Impact of the COVID-19 Pandemic on Breast Cancer Care. JCO Oncol Pract. Published online March 19, 2021:OP.20.01062. doi:10.1200/OP.20.01062

16. Bakouny Z, Paciotti M, Schmidt AL, Lipsitz SR, Choueiri TK, Trinh Q-D. Cancer Screening Tests and Cancer Diagnoses During the COVID-19 Pandemic. JAMA Oncol. 2021;7(3):458–460. doi:10.1001/jamaoncol.2020.7600

17. Wu J, Bobo S, Henry S, Mills M, Kurian A, Dirbas F. Abstract PS6-32: Impact of COVID-19 on breast cancer care at a Bay Area academic center. Cancer Res. 2021;81(4 Supplement):PS6-PS6-32. doi:10.1158/1538-7445.SABCS20-PS6-32

18. Burki TK. Cancer guidelines during the COVID-19 pandemic. Lancet Oncol. 2020;21(5):629–630. doi:10.1016/S1470-2045(20)30217-5

19. Grivas P, Khaki AR, Wise-Draper TM, et al. Association of clinical factors and recent anticancer therapy with COVID-19 severity among patients with cancer: a report from the COVID-19 and Cancer Consortium. Ann Oncol. Published online March 19, 2021. doi:10.1016/j.annonc.2021.02.024

20. Tassone D, Thompson A, Connell W, et al. Immunosuppression as a risk factor for COVID-19: a meta-analysis. Intern Med J. 2021;51(2):199–205. doi:https://doi.org/10.1111/imj.15142

21. Chen MF, Coronel MT, Pan S, et al. Abstract S11-02: Factors associated with developing COVID-19 among cancer patients in New York City. Clin Cancer Res. 2021;27(6 Supplement):S11-S11-02. doi:10.1158/1557-3265.COVID-19-21-S11-02

22. Riera R, Bagattini ÂM, Pacheco RL, Pachito DV, Roitberg F, Ilbawi A. Delays and Disruptions in Cancer Health Care Due to COVID-19 Pandemic: Systematic Review. JCO Glob Oncol. 2021;7:311–323. doi:10.1200/go.20.00639

23. Cone EB, Marchese M, Paciotti M, et al. Assessment of Time-to-Treatment Initiation and Survival in a Cohort of Patients With Common Cancers. JAMA Netw Open. 2020;3(12):e2030072–e2030072. doi:10.1001/jamanetworkopen.2020.30072

24. Hanna TP, King WD, Thibodeau S, et al. Mortality due to cancer treatment delay: systematic review and meta-analysis. BMJ. Published online November 4, 2020:m4087. doi:10.1136/bmj.m4087

25. Khorana AA, Tullio K, Elson P, et al. Time to initial cancer treatment in the United States and association with survival over time: An observational study. PLOS ONE. 2019;14(3):e0213209. doi:10.1371/journal.pone.0213209

26. Bleicher RJ, Ruth K, Sigurdson ER, et al. Time to Surgery and Breast Cancer Survival in the United States. JAMA Oncol. 2016;2(3):330–339. doi:10.1001/jamaoncol.2015.4508

27. Jung SY, Sereika SM, Linkov F, Brufsky A, Weissfeld JL, Rosenzweig M. The effect of delays in treatment for breast cancer metastasis on survival. Breast Cancer Res Treat. 2011;130(3):953–965. doi:10.1007/s10549-011-1662-4

28. Takvorian SU, Oganisian A, Mamtani R, et al. Association of Medicaid Expansion Under the Affordable Care Act With Insurance Status, Cancer Stage, and Timely Treatment Among Patients With Breast, Colon, and Lung Cancer. JAMA Netw Open. 2020;3(2):e1921653–e1921653. doi:10.1001/jamanetworkopen.2019.21653

29. Adamson BJS, Cohen AB, Estevez M, et al. Affordable Care Act (ACA) Medicaid expansion impact on racial disparities in time to cancer treatment. J Clin Oncol. 2019;37(18_suppl):LBA1-LBA1. doi:10.1200/JCO.2019.37.18_suppl.LBA1

30. Webb Hooper M, Nápoles AM, Pérez-Stable EJ. COVID-19 and Racial/Ethnic Disparities. JAMA. 2020;323(24):2466. doi:10.1001/jama.2020.8598

31. Yancy CW. COVID-19 and African Americans. JAMA. 2020;323(19):1891. doi:10.1001/jama.2020.6548

32. Esai Selvan M. Risk factors for death from COVID-19. Nat Rev Immunol. 2020;20(7):407–407. doi:10.1038/s41577-020-0351-0

33. Mueller AL, McNamara MS, Sinclair DA. Why does COVID-19 disproportionately affect older people? Aging. 2020;12(10):9959–9981. doi:10.18632/aging.103344

34. Wang Q, Berger NA, Xu R. Analyses of Risk, Racial Disparity, and Outcomes Among US Patients With Cancer and COVID-19 Infection. JAMA Oncol. 2021;7(2):220–227. doi:10.1001/jamaoncol.2020.6178

35. Holmes JA, Chen RC. Racial Disparities in Time From Diagnosis to Treatment for Stage I Non–Small Cell Lung Cancer. JNCI Cancer Spectr. 2018;2(pky007). doi:10.1093/jncics/pky007

36. Ailawadhi S, Parikh K, Abouzaid S, et al. Racial disparities in treatment patterns and outcomes among patients with multiple myeloma: a SEER-Medicare analysis. Blood Adv. 2019;3(20):2986–2994. doi:10.1182/bloodadvances.2019000308

37. Frankenfeld CL, Menon N, Leslie TF. Racial disparities in colorectal cancer time-to-treatment and survival time in relation to diagnosing hospital cancer-related diagnostic and treatment capabilities. Cancer Epidemiol. 2020;65:101684. doi:10.1016/j.canep.2020.101684

38. Blom EF, ten Haaf K, Arenberg DA, de Koning HJ. Disparities in Receiving Guideline-Concordant Treatment for Lung Cancer in the United States. Ann Am Thorac Soc. 2019;17(2):186–194. doi:10.1513/AnnalsATS.201901-094OC

39. Earle CC, Venditti LN, Neumann PJ, et al. Who Gets Chemotherapy for Metastatic Lung Cancer?(*). Chest. 2000;117(5):1239–1239.

40. Khanal N, Upadhyay S, Dahal S, Bhatt VR, Silberstein PT. Systemic therapy in stage IV pancreatic cancer: a population-based analysis using the National Cancer Data Base. Ther Adv Med Oncol. 2015;7(4):198–205. doi:10.1177/1758834015579313

41. Ma X, Long L, Moon S, Adamson BJS, Baxi SS. Comparison of Population Characteristics in Real-World Clinical Oncology Databases in the US: Flatiron Health, SEER, and NPCR. medRxiv. Published online May 30, 2020:2020.03.16.20037143. doi:10.1101/2020.03.16.20037143

42. Birnbaum B, Nussbaum N, Seidl-Rathkopf K, et al. Model-assisted cohort selection with bias analysis for generating large-scale cohorts from the EHR for oncology research. ArXiv200109765 Cs. Published online January 13, 2020. Accessed April 23, 2021. http://arxiv.org/abs/2001.09765

43. Ettinger DS, Wood DE, Aggarwal C, et al. NCCN Guidelines Insights: Non–Small Cell Lung Cancer, Version 1.2020: Featured Updates to the NCCN Guidelines. J Natl Compr Canc Netw. 2019;17(12):1464–1472. doi:10.6004/jnccn.2019.0059

44. Flaig TW, Spiess PE, Agarwal N, et al. Bladder Cancer, Version 3.2020, NCCN Clinical Practice Guidelines in Oncology. J Natl Compr Canc Netw. 2020;18(3):329–354. doi:10.6004/jnccn.2020.0011

45. Gradishar WJ, Anderson BO, Abraham J, et al. Breast Cancer, Version 3.2020, NCCN Clinical Practice Guidelines in Oncology. J Natl Compr Canc Netw. 2020;18(4):452–478. doi:10.6004/jnccn.2020.0016

46. Mohler JL, Antonarakis ES, Armstrong AJ, et al. Prostate Cancer, Version 2.2019, NCCN Clinical Practice Guidelines in Oncology. J Natl Compr Canc Netw. 2019;17(5):479–505. doi:10.6004/jnccn.2019.0023

47. Kasim RM, Raudenbush SW. Application of Gibbs Sampling to Nested Variance Components Models with Heterogeneous Within-Group Variance. J Educ Behav Stat. 1998;23(2):93–116. doi:10.2307/1165316

48. Austin PC. Balance diagnostics for comparing the distribution of baseline covariates between treatment groups in propensity-score matched samples. Stat Med. 2009;28(25):3083–3107. doi:10.1002/sim.3697

49. Roderick J. A. Little, Donald B. Rubin. Statistical Analysis with Missing Data, 3rd Edition. Wiley Accessed April 23, 2021. https://www.wiley.com/en-us/Statistical+Analysis+with+Missing+Data%2C+3rd+Edition-p-9780470526798

50. Buuren S van, Groothuis-Oudshoorn K. mice: Multivariate Imputation by Chained Equations in R. J Stat Softw. 2011;45(1):1–67. doi:10.18637/jss.v045.i03

51. Borgan Ø. Modeling Survival Data: Extending the Cox Model. Terry M. Therneau and Patricia M. Grambsch, Springer-Verlag, New York, 2000. No. of pages: xiii + 350. Price: $69.95. ISBN 0-387-98784-3. Stat Med. 2001;20(13):2053–2054. doi:https://doi.org/10.1002/sim.956

52. Sjölander A. Regression standardization with the R package stdReg. Eur J Epidemiol. 2016;31(6):563–574. doi:10.1007/s10654-016-0157-3

53. Hassett MJ, Uno H, Cronin AM, Carroll NM, Hornbrook MC, Ritzwoller DP. Comparing Survival After Recurrent vs De Novo Stage IV Advanced Breast, Lung, and Colorectal Cancer. JNCI Cancer Spectr. 2018;2(pky024). doi:10.1093/jncics/pky024

54. Yardley DA, Kaufman PA, Brufsky A, et al. Treatment patterns and clinical outcomes for patients with de novo versus recurrent HER2-positive metastatic breast cancer. Breast Cancer Res Treat. 2014;145(3):725–734. doi:10.1007/s10549-014-2916-8

55. Pottegård A, Kurz X, Moore N, Christiansen CF, Klungel O. Considerations for pharmacoepidemiological analyses in the SARS-CoV-2 pandemic. Pharmacoepidemiol Drug Saf. 2020;29(8):825–831. doi:https://doi.org/10.1002/pds.5029

56. Webster-Clark M. Ways COVID-19 may impact unrelated pharmacoepidemiologic research using routinely collected data. Pharmacoepidemiol Drug Saf. 2021;30(3):400–401. doi:https://doi.org/10.1002/pds.5182

57. Lambe M, Blomqvist P, Bellocco R. Seasonal variation in the diagnosis of cancer: a study based on national cancer registration in Sweden. Br J Cancer. 2003;88(9):1358–1360. doi:10.1038/sj.bjc.6600901

