## Supplementary material for "Impact of the COVID-19 pandemic on treatment patterns for US patients with metastatic solid cancer": PRACTICE Supplemental File

### eTable 1. Stay at home order dates by state/territory

| State | Date | Level |
| --- | --- | --- |
| California | 3/19/2020 | State |
| Illinois | 3/21/2020 | State |
| New Jersey | 3/21/2020 | State |
| New York | 3/22/2020 | State |
| Connecticut | 3/23/2020 | State |
| Louisiana | 3/23/2020 | State |
| Ohio | 3/23/2020 | State |
| Oregon | 3/23/2020 | State |
| Washington | 3/23/2020 | State |
| Delaware | 3/24/2020 | State |
| Indiana | 3/24/2020 | State |
| Massachusetts | 3/24/2020 | State |
| Michigan | 3/24/2020 | State |
| New Mexico | 3/24/2020 | State |
| West Virginia | 3/24/2020 | State |
| Hawaii | 3/25/2020 | State |
| Idaho | 3/25/2020 | State |
| Oklahoma ^a^ | 3/25/2020 | City |
| Vermont | 3/25/2020 | State |
| Wisconsin | 3/25/2020 | State |
| Colorado | 3/26/2020 | State |
| Minnesota | 3/27/2020 | State |
| New Hampshire | 3/27/2020 | State |
| Utah ^b^ | 3/27/2020 | County |
| Alaska | 3/28/2020 | State |
| Montana | 3/28/2020 | State |
| Rhode Island | 3/28/2020 | State |
| Wyoming ^c^ | 3/28/2020 | County |
| Kansas | 3/30/2020 | State |
| Maryland | 3/30/2020 | State |
| North Carolina | 3/30/2020 | State |
| Virginia | 3/30/2020 | State |
| Arizona | 3/31/2020 | State |
| Tennessee | 3/31/2020 | State |
| District of Columbia | 4/1/2020 | State |
| Nevada | 4/1/2020 | State |
| Pennsylvania | 4/1/2020 | State |
| Maine | 4/2/2020 | State |
| Texas | 4/2/2020 | State |
| Florida | 4/3/2020 | State |
| Georgia | 4/3/2020 | State |
| Mississippi  Alabama | 4/3/2020  4/4/2020 | State  State |
| Missouri | 4/6/2020 | State |
| South Carolina | 4/7/2020 | State |
| Kentucky ^d^ |  | None |
| Arkansas |  | None |
| Iowa |  | None |
| Nebraska |  | None |
| North Dakota |  | None |
| South Dakota |  | None |
| ^a^ Oklahoma: initial stay at home/shelter in place orders were given to major cities, beginning with Norman (March 25), Oklahoma City (March 28), and Tulsa (March 28) | | |
| ^b^ Utah: Initial stay at home orders were county-specific, starting with Summit County(March 27), Salt Lake County (March 30), and Davis County(April 1) | | |
| ^c^ Wyoming: Stay at home orders were county-specific, beginning with Jackson County (March 28) | | |
| ^d^ Kentucky: A state-level "healthy at home" recommendation was issued on March 26, but no full stay-at-home orders were issued. | | |

### eFigure 1. CONSORT diagram

|  | Criteria | Patients lost | Patients remaining |
| --- | --- | --- | --- |
|  | Total unique patients |  | 164,188 |
|  | Exclude patients outside of study frame (all patients not diagnosed in Jan-Jul 2019 or Jan-Jul 2020) | 143,927 | 20,261 |
|  | Exclude patients with no structured activity within 90 days of diagnosis | 1,631 | 18,630 |
|  | Exclude patients with <2 observations after diagnoses | 1,275 | 17,355 |
|  | Exclude patients with multiple malignancies (only study cohorts considered) | 66 | 17,289 |
|  | Treatment begins prior to advanced diagnosis | 682 | 16,607 |
|  | Exclude patients on non-NCCN recommended therapy or a therapy that involved a clinical study drug | 344 | 16,263 |
|  | Washout | 2,127 | 14,136 |
| Aim 2 | Excluded patients who were not diagnosed with breast cancer, prostate cancer, urothelial cancer, or NSCLC. | 4,603 | 9,533 |
|  | Excluded patients not treated within 60 days of advanced diagnosis. | 2,812 | 6,721 |
|  | Excluded patients who received targeted therapy | 759 | 5,962 |

^a^ NCCN = National Comprehensive Cancer Network; NSCLC = Non-small cell lung carcinoma

### eTable 2. Therapy classifications

| Cancer | Drug Name | Clinical Study Drug | NCCN Approved | Targeted | Myelosuppresive |
| --- | --- | --- | --- | --- | --- |
| Breast | Abemaciclib,Anastrozole,Fulvestrant | No | Yes | No | Yes |
| Breast | Abemaciclib,Anastrozole,Fulvestrant,Palbociclib | No | Yes | No | Yes |
| Breast | Abemaciclib,Anastrozole,Palbociclib | No | Yes | No | Yes |
| Breast | Abemaciclib,Fulvestrant,Goserelin | No | Yes | No | Yes |
| Breast | Abemaciclib,Fulvestrant,Letrozole,Palbociclib | No | Yes | No | Yes |
| Breast | Abemaciclib,Fulvestrant,Ribociclib | No | Yes | No | Yes |
| Breast | Abemaciclib,Letrozole,Leuprolide,Ribociclib | No | Yes | No | Yes |
| Breast | Abemaciclib,Leuprolide | No | Yes | No | Yes |
| Breast | Abemaciclib,Paclitaxel Protein-Bound | No | Yes | No | Yes |
| Breast | Abemaciclib,Palbociclib | No | Yes | No | Yes |
| Breast | Ado-Trastuzumab Emtansine,Exemestane | No | Yes | Yes |  |
| Breast | Ado-Trastuzumab Emtansine,Letrozole | No | Yes | Yes |  |
| Breast | Ado-Trastuzumab Emtansine,Leuprolide,Tamoxifen | No | Yes | Yes |  |
| Breast | Ado-Trastuzumab Emtansine,Pertuzumab,Trastuzumab-Anns | No | Yes | Yes |  |
| Breast | Alpelisib | No | Yes | Yes |  |
| Breast | Alpelisib,Fulvestrant,Palbociclib | No | Yes | Yes |  |
| Breast | Anastrozole,Carboplatin,Gemcitabine | No | Yes | No | Yes |
| Breast | Anastrozole,Gemcitabine,Paclitaxel | No | Yes | No | Yes |
| Breast | Anastrozole,Goserelin | No | Yes | No | No |
| Breast | Anastrozole,Leuprolide,Palbociclib,Pertuzumab,Trastuzumab-Anns | No | Yes | Yes |  |
| Breast | Anastrozole,Palbociclib,Pertuzumab,Trastuzumab-Anns | No | Yes | Yes |  |
| Breast | Anastrozole,Pertuzumab,Trastuzumab-Anns | No | Yes | Yes |  |
| Breast | Anastrozole,Trastuzumab-Anns | No | Yes | Yes |  |
| Breast | Anastrozole,Triptorelin | No | Yes | No | No |
| Breast | Atezolizumab | No | Yes | Yes |  |
| Breast | Capecitabine,Clinical Study Drug | Yes |  |  |  |
| Breast | Capecitabine,Doxorubicin Pegylated Liposomal | No | Yes | No | Yes |
| Breast | Capecitabine,Fulvestrant | No | Yes | No | Yes |
| Breast | Capecitabine,Letrozole,Oxaliplatin,Palbociclib | No | Yes | No | Yes |
| Breast | Capecitabine,Leuprolide,Ribociclib,Tamoxifen | No | Yes | No | Yes |
| Breast | Capecitabine,Trastuzumab-Anns | No | Yes | Yes |  |
| Breast | Capecitabine,Trastuzumab-Anns,Tucatinib | No | Yes | Yes |  |
| Breast | Capecitabine,Trastuzumab,Tucatinib | No | Yes | Yes |  |
| Breast | Carboplatin,Docetaxel,Gemcitabine,Paclitaxel | No | Yes | No | Yes |
| Breast | Carboplatin,Docetaxel,Leuprolide,Pertuzumab,Trastuzumab-Anns | No | Yes | Yes |  |
| Breast | Carboplatin,Docetaxel,Trastuzumab | No | Yes | Yes |  |
| Breast | Carboplatin,Eribulin,Letrozole | No | Yes | No | Yes |
| Breast | Carboplatin,Gemcitabine,Pembrolizumab | No | Yes | Yes |  |
| Breast | Clinical Study Drug,Fulvestrant,Palbociclib | Yes |  |  |  |
| Breast | Crizotinib,Vinorelbine | No | Yes | No | Yes |
| Breast | Cyclophosphamide,Doxorubicin,Paclitaxel | No | Yes | No | Yes |
| Breast | Cyclophosphamide,Epirubicin | No | Yes | No | Yes |
| Breast | Degarelix | No | No |  |  |
| Breast | Docetaxel,Fulvestrant,Pertuzumab,Trastuzumab | No | Yes | Yes |  |
| Breast | Docetaxel,Letrozole,Paclitaxel,Pertuzumab,Trastuzumab-Anns | No | Yes | Yes |  |
| Breast | Docetaxel,Paclitaxel | No | Yes | No | Yes |
| Breast | Docetaxel,Pertuzumab,Trastuzumab-Qyyp | No | Yes | Yes |  |
| Breast | Doxorubicin Pegylated Liposomal,Fulvestrant | No | Yes | No | Yes |
| Breast | Eribulin,Letrozole | No | Yes | No | Yes |
| Breast | Eribulin,Leuprolide,Tamoxifen | No | Yes | No | Yes |
| Breast | Eribulin,Tamoxifen | No | Yes | No | Yes |
| Breast | Everolimus | No | Yes | No | Yes |
| Breast | Exemestane,Goserelin | No | Yes | No | No |
| Breast | Exemestane,Ribociclib | No | Yes | No | Yes |
| Breast | Fluorouracil,Leucovorin | No | No |  |  |
| Breast | Fulvestrant,Goserelin | No | Yes | No | No |
| Breast | Fulvestrant,Letrozole,Leuprolide | No | Yes | No | No |
| Breast | Fulvestrant,Paclitaxel,Palbociclib | No | Yes | No | Yes |
| Breast | Fulvestrant,Palbociclib,Ribociclib | No | Yes | No | Yes |
| Breast | Fulvestrant,Palbociclib,Tamoxifen | No | Yes | No | Yes |
| Breast | Gemcitabine,Pertuzumab,Trastuzumab | No | Yes | Yes |  |
| Breast | Goserelin,Palbociclib | No | Yes | No | Yes |
| Breast | Goserelin,Pertuzumab,Trastuzumab | No | Yes | Yes |  |
| Breast | Ixabepilone | No | Yes | No | Yes |
| Breast | Letrozole,Leuprolide,Palbociclib,Trastuzumab | No | Yes | Yes |  |
| Breast | Letrozole,Neratinib | No | Yes | Yes |  |
| Breast | Letrozole,Paclitaxel,Pertuzumab,Trastuzumab-Anns | No | Yes | Yes |  |
| Breast | Leuprolide,Palbociclib | No | Yes | No | Yes |
| Breast | Leuprolide,Palbociclib,Tamoxifen | No | Yes | No | Yes |
| Breast | Neratinib | No | Yes | Yes |  |
| Breast | Paclitaxel Protein-Bound,Pembrolizumab,Vinorelbine | No | Yes | Yes |  |
| Breast | Paclitaxel,Palbociclib | No | Yes | No | Yes |
| Breast | Paclitaxel,Ruxolitinib,Trastuzumab | No | Yes | Yes |  |
| Breast | Pertuzumab,Tamoxifen,Trastuzumab | No | Yes | Yes |  |
| Breast | Tucatinib | No | Yes | Yes |  |
| Breast | Abemaciclib | No | Yes | No | Yes |
| Breast | Abemaciclib,Anastrozole | No | Yes | No | Yes |
| Breast | Abemaciclib,Anastrozole,Goserelin | No | Yes | No | Yes |
| Breast | Abemaciclib,Carboplatin,Docetaxel,Fulvestrant | No | Yes | No | Yes |
| Breast | Abemaciclib,Cyclophosphamide,Doxorubicin,Letrozole | No | Yes | No | Yes |
| Breast | Abemaciclib,Exemestane | No | Yes | No | Yes |
| Breast | Abemaciclib,Fulvestrant | No | Yes | No | Yes |
| Breast | Abemaciclib,Fulvestrant,Letrozole | No | Yes | No | Yes |
| Breast | Abemaciclib,Fulvestrant,Leuprolide | No | Yes | No | Yes |
| Breast | Abemaciclib,Fulvestrant,Paclitaxel Protein-Bound | No | Yes | No | Yes |
| Breast | Abemaciclib,Fulvestrant,Palbociclib | No | Yes | No | Yes |
| Breast | Abemaciclib,Goserelin,Letrozole | No | Yes | No | Yes |
| Breast | Abemaciclib,Letrozole | No | Yes | No | Yes |
| Breast | Abemaciclib,Letrozole,Leuprolide | No | Yes | No | Yes |
| Breast | Abemaciclib,Letrozole,Palbociclib | No | Yes | No | Yes |
| Breast | Abemaciclib,Leuprolide,Tamoxifen | No | Yes | No | Yes |
| Breast | Abemaciclib,Ribociclib | No | Yes | No | Yes |
| Breast | Abemaciclib,Tamoxifen | No | Yes | No | Yes |
| Breast | Ado-Trastuzumab Emtansine | No | Yes | Yes |  |
| Breast | Ado-Trastuzumab Emtansine,Exemestane,Goserelin | No | Yes | Yes |  |
| Breast | Ado-Trastuzumab Emtansine,Paclitaxel Protein-Bound | No | Yes | Yes |  |
| Breast | Ado-Trastuzumab Emtansine,Pertuzumab | No | Yes | Yes |  |
| Breast | Ado-Trastuzumab Emtansine,Pertuzumab/Trastuzumab/Hyaluronidase-Zzxf | No | Yes | Yes |  |
| Breast | Alpelisib,Carboplatin,Fulvestrant,Gemcitabine | No | Yes | Yes |  |
| Breast | Alpelisib,Fulvestrant | No | Yes | Yes |  |
| Breast | Anastrozole | No | Yes | No | No |
| Breast | Anastrozole,Capecitabine | No | Yes | No | Yes |
| Breast | Anastrozole,Clinical Study Drug | Yes |  |  |  |
| Breast | Anastrozole,Clinical Study Drug,Ribociclib | Yes |  |  |  |
| Breast | Anastrozole,Cyclophosphamide,Doxorubicin | No | Yes | No | Yes |
| Breast | Anastrozole,Docetaxel | No | Yes | No | Yes |
| Breast | Anastrozole,Docetaxel,Pertuzumab,Trastuzumab | No | Yes | Yes |  |
| Breast | Anastrozole,Fulvestrant | No | Yes | No | No |
| Breast | Anastrozole,Fulvestrant,Goserelin | No | Yes | No | No |
| Breast | Anastrozole,Fulvestrant,Palbociclib | No | Yes | No | Yes |
| Breast | Anastrozole,Fulvestrant,Trastuzumab | No | Yes | Yes |  |
| Breast | Anastrozole,Goserelin,Palbociclib | No | Yes | No | Yes |
| Breast | Anastrozole,Leuprolide | No | Yes | No | No |
| Breast | Anastrozole,Leuprolide,Palbociclib | No | Yes | No | Yes |
| Breast | Anastrozole,Paclitaxel Protein-Bound,Pembrolizumab | No | Yes | Yes |  |
| Breast | Anastrozole,Paclitaxel,Pertuzumab,Trastuzumab-Anns | No | Yes | Yes |  |
| Breast | Anastrozole,Paclitaxel,Trastuzumab | No | Yes | Yes |  |
| Breast | Anastrozole,Palbociclib | No | Yes | No | Yes |
| Breast | Anastrozole,Palbociclib,Pertuzumab,Trastuzumab | No | Yes | Yes |  |
| Breast | Anastrozole,Palbociclib,Ribociclib | No | Yes | No | Yes |
| Breast | Anastrozole,Pertuzumab,Trastuzumab | No | Yes | Yes |  |
| Breast | Anastrozole,Ribociclib | No | Yes | No | Yes |
| Breast | Anastrozole,Trastuzumab | No | Yes | Yes |  |
| Breast | Atezolizumab,Capecitabine | No | Yes | Yes |  |
| Breast | Atezolizumab,Capecitabine,Paclitaxel Protein-Bound | No | Yes | Yes |  |
| Breast | Atezolizumab,Carboplatin,Paclitaxel,Paclitaxel Protein-Bound | No | Yes | Yes |  |
| Breast | Atezolizumab,Cyclophosphamide,Doxorubicin,Paclitaxel,Paclitaxel Protein-Bound | No | Yes | Yes |  |
| Breast | Atezolizumab,Docetaxel,Paclitaxel,Paclitaxel Protein-Bound | No | Yes | Yes |  |
| Breast | Atezolizumab,Fulvestrant,Paclitaxel Protein-Bound | No | Yes | Yes |  |
| Breast | Atezolizumab,Paclitaxel Protein-Bound | No | Yes | Yes |  |
| Breast | Atezolizumab,Paclitaxel Protein-Bound,Pembrolizumab | No | Yes | Yes |  |
| Breast | Atezolizumab,Paclitaxel Protein-Bound,Samarium Sm 153 Lexidronam | No | Yes | Yes |  |
| Breast | Atezolizumab,Paclitaxel Protein-Bound,Tamoxifen | No | Yes | Yes |  |
| Breast | Atezolizumab,Paclitaxel,Paclitaxel Protein-Bound | No | Yes | Yes |  |
| Breast | Bevacizumab | No | No |  |  |
| Breast | Bevacizumab,Carboplatin | No | Yes | No | Yes |
| Breast | Bevacizumab,Carboplatin,Paclitaxel | No | Yes | No | Yes |
| Breast | Bevacizumab,Docetaxel | No | Yes | No | Yes |
| Breast | Bevacizumab,Paclitaxel Protein-Bound | No | Yes | No | Yes |
| Breast | Bevacizumab,Paclitaxel,Paclitaxel Protein-Bound | No | Yes | No | Yes |
| Breast | Bicalutamide,Cisplatin | No | Yes | No | Yes |
| Breast | Bortezomib | No | No |  |  |
| Breast | Calaspargase Pegol-Mknl,Trastuzumab | No | Yes | Yes |  |
| Breast | Capecitabine | No | Yes | No | Yes |
| Breast | Capecitabine,Carboplatin,Gemcitabine | No | Yes | Yes |  |
| Breast | Capecitabine,Carboplatin,Gemcitabine,Pembrolizumab | No | Yes | Yes |  |
| Breast | Capecitabine,Docetaxel | No | Yes | No | Yes |
| Breast | Capecitabine,Docetaxel,Tamoxifen | No | Yes | No | Yes |
| Breast | Capecitabine,Goserelin,Tamoxifen | No | Yes | No | Yes |
| Breast | Capecitabine,Lapatinib | No | Yes | Yes |  |
| Breast | Capecitabine,Letrozole | No | Yes | No | Yes |
| Breast | Capecitabine,Letrozole,Oxaliplatin | No | Yes | No | Yes |
| Breast | Capecitabine,Paclitaxel Protein-Bound | No | Yes | No | Yes |
| Breast | Capecitabine,Trastuzumab | No | Yes | Yes |  |
| Breast | Capecitabine,Uridine | No | Yes | No | Yes |
| Breast | Carboplatin | No | Yes | No | Yes |
| Breast | Carboplatin,Cisplatin,Paclitaxel | No | Yes | No | Yes |
| Breast | Carboplatin,Cyclophosphamide,Docetaxel,Doxorubicin | No | Yes | No | Yes |
| Breast | Carboplatin,Docetaxel | No | Yes | No | Yes |
| Breast | Carboplatin,Docetaxel,Goserelin,Pertuzumab,Trastuzumab-Anns | No | Yes | Yes |  |
| Breast | Carboplatin,Docetaxel,Pertuzumab,Tamoxifen,Trastuzumab-Anns | No | Yes | Yes |  |
| Breast | Carboplatin,Docetaxel,Pertuzumab,Trastuzumab | No | Yes | Yes |  |
| Breast | Carboplatin,Docetaxel,Pertuzumab,Trastuzumab-Anns | No | Yes | Yes |  |
| Breast | Carboplatin,Docetaxel,Pertuzumab,Trastuzumab-Dkst | No | Yes | Yes |  |
| Breast | Carboplatin,Docetaxel,Pertuzumab,Trastuzumab-Pkrb | No | Yes | Yes |  |
| Breast | Carboplatin,Docetaxel,Pertuzumab,Trastuzumab-Qyyp | No | Yes | Yes |  |
| Breast | Carboplatin,Eribulin,Gemcitabine | No | Yes | No | Yes |
| Breast | Carboplatin,Gemcitabine | No | Yes | No | Yes |
| Breast | Carboplatin,Gemcitabine,Paclitaxel | No | Yes | No | Yes |
| Breast | Carboplatin,Gemcitabine,Pertuzumab,Trastuzumab/Hyaluronidase-Oysk | No | Yes | Yes |  |
| Breast | Carboplatin,Gemcitabine,Trastuzumab | No | Yes | Yes |  |
| Breast | Carboplatin,Letrozole,Paclitaxel | No | Yes | No | Yes |
| Breast | Carboplatin,Letrozole,Paclitaxel,Paclitaxel Protein-Bound | No | Yes | No | Yes |
| Breast | Carboplatin,Letrozole,Paclitaxel,Palbociclib | No | Yes | No | Yes |
| Breast | Carboplatin,Paclitaxel | No | Yes | No | Yes |
| Breast | Carboplatin,Paclitaxel Protein-Bound | No | Yes | No | Yes |
| Breast | Carboplatin,Paclitaxel,Trastuzumab-Anns | No | Yes | Yes |  |
| Breast | Carboplatin,Talazoparib | No | Yes | Yes |  |
| Breast | Cisplatin | No | Yes | No | Yes |
| Breast | Cisplatin,Etoposide | No | Yes | No | Yes |
| Breast | Cisplatin,Gemcitabine | No | Yes | No | Yes |
| Breast | Cisplatin,Letrozole | No | Yes | No | Yes |
| Breast | Clinical Study Drug | Yes |  |  |  |
| Breast | Clinical Study Drug,Docetaxel | Yes |  |  |  |
| Breast | Clinical Study Drug,Fulvestrant | Yes |  |  |  |
| Breast | Clinical Study Drug,Fulvestrant,Ribociclib | Yes |  |  |  |
| Breast | Clinical Study Drug,Letrozole | Yes |  |  |  |
| Breast | Clinical Study Drug,Letrozole,Ribociclib | Yes |  |  |  |
| Breast | Cyclophosphamide,Docetaxel | No | Yes | No | Yes |
| Breast | Cyclophosphamide,Docetaxel,Doxorubicin | No | Yes | No | Yes |
| Breast | Cyclophosphamide,Docetaxel,Doxorubicin,Paclitaxel | No | Yes | No | Yes |
| Breast | Cyclophosphamide,Docetaxel,Doxorubicin,Pertuzumab,Trastuzumab | No | Yes | Yes |  |
| Breast | Cyclophosphamide,Docetaxel,Fulvestrant,Paclitaxel,Pertuzumab,Trastuzumab-Anns | No | Yes | Yes |  |
| Breast | Cyclophosphamide,Docetaxel,Letrozole | No | Yes | No | Yes |
| Breast | Cyclophosphamide,Docetaxel,Paclitaxel Protein-Bound | No | Yes | No | Yes |
| Breast | Cyclophosphamide,Doxorubicin | No | Yes | No | Yes |
| Breast | Cyclophosphamide,Doxorubicin,Letrozole,Palbociclib | No | Yes | No | Yes |
| Breast | Cyclophosphamide,Doxorubicin,Leuprolide | No | Yes | No | Yes |
| Breast | Cyclophosphamide,Doxorubicin,Leuprolide,Tamoxifen | No | Yes | No | Yes |
| Breast | Cyclophosphamide,Doxorubicin,Pembrolizumab | No | Yes | Yes |  |
| Breast | Cyclophosphamide,Fluorouracil,Methotrexate | No | Yes | No | Yes |
| Breast | Docetaxel | No | Yes | No | Yes |
| Breast | Docetaxel,Fluorouracil,Leucovorin,Oxaliplatin | No | Yes | No | Yes |
| Breast | Docetaxel,Fulvestrant,Palbociclib | No | Yes | No | Yes |
| Breast | Docetaxel,Gemcitabine | No | Yes | No | Yes |
| Breast | Docetaxel,Letrozole,Paclitaxel,Pertuzumab,Trastuzumab | No | Yes | Yes |  |
| Breast | Docetaxel,Letrozole,Pertuzumab,Trastuzumab-Anns | No | Yes | Yes |  |
| Breast | Docetaxel,Letrozole,Pertuzumab,Trastuzumab-Pkrb | No | Yes | Yes |  |
| Breast | Docetaxel,Leuprolide,Pertuzumab,Trastuzumab | No | Yes | Yes |  |
| Breast | Docetaxel,Leuprolide,Pertuzumab,Trastuzumab-Anns | No | Yes | Yes |  |
| Breast | Docetaxel,Paclitaxel Protein-Bound,Pertuzumab,Trastuzumab-Dkst | No | Yes | Yes |  |
| Breast | Docetaxel,Paclitaxel,Pertuzumab,Trastuzumab | No | Yes | Yes |  |
| Breast | Docetaxel,Paclitaxel,Trastuzumab-Anns | No | Yes | Yes |  |
| Breast | Docetaxel,Pertuzumab,Tamoxifen,Trastuzumab,Triptorelin | No | Yes | Yes |  |
| Breast | Docetaxel,Pertuzumab,Trastuzumab | No | Yes | Yes |  |
| Breast | Docetaxel,Pertuzumab,Trastuzumab-Anns | No | Yes | Yes |  |
| Breast | Docetaxel,Pertuzumab,Trastuzumab-Dkst | No | Yes | Yes |  |
| Breast | Docetaxel,Pertuzumab,Trastuzumab-Pkrb | No | Yes | Yes |  |
| Breast | Docetaxel,Pertuzumab,Trastuzumab,Trastuzumab-Qyyp | No | Yes | Yes |  |
| Breast | Doxorubicin | No | Yes | No | Yes |
| Breast | Doxorubicin Pegylated Liposomal | No | Yes | No | Yes |
| Breast | Doxorubicin Pegylated Liposomal,Exemestane | No | Yes | No | Yes |
| Breast | Doxorubicin Pegylated Liposomal,Trastuzumab-Dkst | No | Yes | Yes |  |
| Breast | Doxorubicin,Paclitaxel,Trastuzumab | No | Yes | Yes |  |
| Breast | Eribulin | No | Yes | No | Yes |
| Breast | Eribulin,Leuprolide | No | Yes | No | Yes |
| Breast | Everolimus,Exemestane | No | Yes | No | Yes |
| Breast | Exemestane | No | Yes | No | No |
| Breast | Exemestane,Fulvestrant | No | Yes | No | No |
| Breast | Exemestane,Fulvestrant,Paclitaxel,Pertuzumab,Trastuzumab | No | Yes | Yes |  |
| Breast | Exemestane,Fulvestrant,Palbociclib | No | Yes | No | Yes |
| Breast | Exemestane,Goserelin,Palbociclib | No | Yes | No | Yes |
| Breast | Exemestane,Goserelin,Ribociclib | No | Yes | No | Yes |
| Breast | Exemestane,Leuprolide,Pertuzumab,Trastuzumab-Pkrb | No | Yes | Yes |  |
| Breast | Exemestane,Palbociclib | No | Yes | No | Yes |
| Breast | Exemestane,Trastuzumab | No | Yes | Yes |  |
| Breast | Fam-Trastuzumab Deruxtecan-Nxki | No | Yes | Yes |  |
| Breast | Fulvestrant | No | Yes | No | No |
| Breast | Fulvestrant,Goserelin,Palbociclib | No | Yes | No | Yes |
| Breast | Fulvestrant,Letrozole | No | Yes | No | No |
| Breast | Fulvestrant,Letrozole,Palbociclib | No | Yes | No | Yes |
| Breast | Fulvestrant,Leuprolide | No | Yes | No | No |
| Breast | Fulvestrant,Leuprolide,Palbociclib | No | Yes | No | Yes |
| Breast | Fulvestrant,Methotrexate | No | Yes | No | Yes |
| Breast | Fulvestrant,Paclitaxel | No | Yes | No | Yes |
| Breast | Fulvestrant,Paclitaxel,Paclitaxel Protein-Bound | No | Yes | No | Yes |
| Breast | Fulvestrant,Palbociclib | No | Yes | No | Yes |
| Breast | Fulvestrant,Pertuzumab,Trastuzumab | No | Yes | Yes |  |
| Breast | Fulvestrant,Ribociclib | No | Yes | No | Yes |
| Breast | Fulvestrant,Tamoxifen | No | Yes | No | No |
| Breast | Fulvestrant,Trastuzumab | No | Yes | Yes |  |
| Breast | Gemcitabine | No | Yes | No | Yes |
| Breast | Gemcitabine,Paclitaxel | No | Yes | No | Yes |
| Breast | Gemcitabine,Pembrolizumab | No | Yes | Yes |  |
| Breast | Gemcitabine,Tamoxifen | No | Yes | No | Yes |
| Breast | Gemcitabine,Vinorelbine | No | Yes | No | Yes |
| Breast | Goserelin | No | Yes | No | No |
| Breast | Goserelin,Letrozole,Palbociclib | No | Yes | No | Yes |
| Breast | Goserelin,Letrozole,Pertuzumab,Trastuzumab | No | Yes | Yes |  |
| Breast | Goserelin,Letrozole,Ribociclib | No | Yes | No | Yes |
| Breast | Goserelin,Palbociclib,Tamoxifen | No | Yes | No | Yes |
| Breast | Goserelin,Ribociclib,Tamoxifen | No | Yes | No | Yes |
| Breast | Goserelin,Tamoxifen | No | Yes | No | No |
| Breast | Hydroxyurea,Paclitaxel | No | Yes | No | Yes |
| Breast | Lapatinib | No | Yes | Yes |  |
| Breast | Lapatinib,Letrozole | No | Yes | Yes |  |
| Breast | Lapatinib,Trastuzumab | No | Yes | Yes |  |
| Breast | Letrozole | No | Yes | No | No |
| Breast | Letrozole,Leuprolide | No | Yes | No | No |
| Breast | Letrozole,Leuprolide,Palbociclib | No | Yes | No | Yes |
| Breast | Letrozole,Leuprolide,Ribociclib | No | Yes | No | Yes |
| Breast | Letrozole,Leuprolide,Trastuzumab | No | Yes | Yes |  |
| Breast | Letrozole,Paclitaxel,Pertuzumab,Trastuzumab | No | Yes | Yes |  |
| Breast | Letrozole,Paclitaxel,Trastuzumab-Anns | No | Yes | Yes |  |
| Breast | Letrozole,Palbociclib | No | Yes | No | Yes |
| Breast | Letrozole,Palbociclib,Ribociclib | No | Yes | No | Yes |
| Breast | Letrozole,Palbociclib,Rituximab | No | Yes | No | Yes |
| Breast | Letrozole,Palbociclib,Trastuzumab | No | Yes | Yes |  |
| Breast | Letrozole,Pertuzumab,Trastuzumab | No | Yes | Yes |  |
| Breast | Letrozole,Pertuzumab,Trastuzumab-Anns | No | Yes | Yes |  |
| Breast | Letrozole,Ribociclib | No | Yes | No | Yes |
| Breast | Letrozole,Ribociclib,Trastuzumab | No | Yes | Yes |  |
| Breast | Letrozole,Trastuzumab | No | Yes | Yes |  |
| Breast | Letrozole,Trastuzumab-Anns | No | Yes | Yes |  |
| Breast | Leucovorin | No | No |  |  |
| Breast | Leucovorin,Methotrexate,Paclitaxel | No | Yes | No | Yes |
| Breast | Leuprolide | No | Yes | No | No |
| Breast | Leuprolide,Niraparib,Pembrolizumab | No | Yes | Yes |  |
| Breast | Leuprolide,Paclitaxel,Pertuzumab,Trastuzumab | No | Yes | Yes |  |
| Breast | Leuprolide,Pertuzumab,Tamoxifen,Trastuzumab-Anns | No | Yes | Yes |  |
| Breast | Leuprolide,Pertuzumab,Trastuzumab-Anns | No | Yes | Yes |  |
| Breast | Leuprolide,Tamoxifen | No | Yes | No | No |
| Breast | Methotrexate | No | Yes | No | Yes |
| Breast | Olaparib | No | Yes | Yes |  |
| Breast | Paclitaxel | No | Yes | No | Yes |
| Breast | Paclitaxel Protein-Bound | No | Yes | No | Yes |
| Breast | Paclitaxel Protein-Bound,Pertuzumab,Trastuzumab | No | Yes | Yes |  |
| Breast | Paclitaxel Protein-Bound,Tamoxifen | No | Yes | No | Yes |
| Breast | Paclitaxel,Paclitaxel Protein-Bound | No | Yes | No | Yes |
| Breast | Paclitaxel,Pertuzumab,Tamoxifen,Trastuzumab | No | Yes | Yes |  |
| Breast | Paclitaxel,Pertuzumab,Trastuzumab | No | Yes | Yes |  |
| Breast | Paclitaxel,Pertuzumab,Trastuzumab-Anns | No | Yes | Yes |  |
| Breast | Paclitaxel,Pertuzumab,Trastuzumab-Pkrb | No | Yes | Yes |  |
| Breast | Paclitaxel,Pertuzumab,Trastuzumab-Qyyp | No | Yes | Yes |  |
| Breast | Paclitaxel,Trastuzumab | No | Yes | Yes |  |
| Breast | Paclitaxel,Trastuzumab-Anns | No | Yes | Yes |  |
| Breast | Palbociclib | No | Yes | No | Yes |
| Breast | Palbociclib,Tamoxifen | No | Yes | No | Yes |
| Breast | Pembrolizumab | No | Yes | Yes |  |
| Breast | Pertuzumab,Tamoxifen,Trastuzumab-Anns | No | Yes | Yes |  |
| Breast | Pertuzumab,Trastuzumab | No | Yes | Yes |  |
| Breast | Pertuzumab,Trastuzumab-Anns | No | Yes | Yes |  |
| Breast | Pertuzumab,Trastuzumab,Vinorelbine | No | Yes | Yes |  |
| Breast | Ribociclib | No | Yes | No | Yes |
| Breast | Ribociclib,Tamoxifen | No | Yes | No | Yes |
| Breast | Sacituzumab Govitecan-Hziy | No | Yes | No | Yes |
| Breast | Talazoparib | No | Yes | Yes |  |
| Breast | Tamoxifen | No | Yes | No | No |
| Breast | Tamoxifen,Trastuzumab | No | Yes | Yes |  |
| Breast | Trastuzumab | No | Yes | Yes |  |
| Breast | Trastuzumab-Anns | No | Yes | Yes |  |
| Breast | Vinorelbine | No | Yes | No | Yes |
| NSCLC | Abemaciclib,Fulvestrant | Yes |  |  |  |
| NSCLC | Abiraterone | Yes |  |  |  |
| NSCLC | Abiraterone,Carboplatin,Pembrolizumab,Pemetrexed | Yes |  |  |  |
| NSCLC | Abiraterone,Carboplatin,Pemetrexed | Yes |  |  |  |
| NSCLC | Abiraterone,Dabrafenib,Trametinib | Yes |  |  |  |
| NSCLC | Abiraterone,Leuprolide | Yes |  |  |  |
| NSCLC | Abiraterone,Pembrolizumab,Pemetrexed | Yes |  |  |  |
| NSCLC | Ado-Trastuzumab Emtansine | Yes |  |  |  |
| NSCLC | Afatinib | Yes |  |  |  |
| NSCLC | Afatinib,Bevacizumab | Yes |  |  |  |
| NSCLC | Afatinib,Letrozole | Yes |  |  |  |
| NSCLC | Afatinib,Leuprolide | Yes |  |  |  |
| NSCLC | Alectinib | Yes |  |  |  |
| NSCLC | Alectinib,Carboplatin,Pemetrexed | Yes |  |  |  |
| NSCLC | Alectinib,Selpercatinib | Yes |  |  |  |
| NSCLC | Alemtuzumab | Yes |  |  |  |
| NSCLC | Anastrozole | Yes |  |  |  |
| NSCLC | Anastrozole,Carboplatin,Paclitaxel | Yes |  |  |  |
| NSCLC | Anastrozole,Carboplatin,Paclitaxel,Pembrolizumab,Pemetrexed | Yes |  |  |  |
| NSCLC | Anastrozole,Carboplatin,Pembrolizumab,Pemetrexed | Yes |  |  |  |
| NSCLC | Anastrozole,Carboplatin,Pemetrexed | Yes |  |  |  |
| NSCLC | Anastrozole,Cisplatin,Pemetrexed | Yes |  |  |  |
| NSCLC | Anastrozole,Letrozole | Yes |  |  |  |
| NSCLC | Anastrozole,Nivolumab | Yes |  |  |  |
| NSCLC | Anastrozole,Osimertinib | Yes |  |  |  |
| NSCLC | Anastrozole,Pembrolizumab | Yes |  |  |  |
| NSCLC | Atezolizumab | Yes |  |  |  |
| NSCLC | Atezolizumab,Bevacizumab | Yes |  |  |  |
| NSCLC | Atezolizumab,Bevacizumab,Carboplatin,Paclitaxel | Yes |  |  |  |
| NSCLC | Atezolizumab,Bevacizumab,Carboplatin,Paclitaxel Protein-Bound | Yes |  |  |  |
| NSCLC | Atezolizumab,Bevacizumab-Awwb,Carboplatin,Paclitaxel | Yes |  |  |  |
| NSCLC | Atezolizumab,Bevacizumab-Awwb,Carboplatin,Paclitaxel Protein-Bound | Yes |  |  |  |
| NSCLC | Atezolizumab,Bevacizumab-Awwb,Carboplatin,Paclitaxel,Rituximab-Pvvr | Yes |  |  |  |
| NSCLC | Atezolizumab,Bevacizumab-Bvzr,Carboplatin,Paclitaxel | Yes |  |  |  |
| NSCLC | Atezolizumab,Bevacizumab-Bvzr,Carboplatin,Paclitaxel Protein-Bound | Yes |  |  |  |
| NSCLC | Atezolizumab,Carboplatin,Etoposide | Yes |  |  |  |
| NSCLC | Atezolizumab,Carboplatin,Etoposide,Paclitaxel Protein-Bound,Pembrolizumab | Yes |  |  |  |
| NSCLC | Atezolizumab,Carboplatin,Etoposide,Paclitaxel,Pembrolizumab | Yes |  |  |  |
| NSCLC | Atezolizumab,Carboplatin,Paclitaxel | Yes |  |  |  |
| NSCLC | Atezolizumab,Carboplatin,Paclitaxel Protein-Bound | Yes |  |  |  |
| NSCLC | Atezolizumab,Carboplatin,Paclitaxel,Pembrolizumab | Yes |  |  |  |
| NSCLC | Atezolizumab,Carboplatin,Pemetrexed | Yes |  |  |  |
| NSCLC | Atezolizumab,Paclitaxel Protein-Bound,Tamoxifen | Yes |  |  |  |
| NSCLC | Azacitidine | Yes |  |  |  |
| NSCLC | Azacitidine,Venetoclax | Yes |  |  |  |
| NSCLC | Bevacizumab | Yes |  |  |  |
| NSCLC | Bevacizumab,Carboplatin | Yes |  |  |  |
| NSCLC | Bevacizumab,Carboplatin,Gemcitabine | Yes |  |  |  |
| NSCLC | Bevacizumab,Carboplatin,Letrozole,Pemetrexed | Yes |  |  |  |
| NSCLC | Bevacizumab,Carboplatin,Paclitaxel | Yes |  |  |  |
| NSCLC | Bevacizumab,Carboplatin,Paclitaxel Protein-Bound | Yes |  |  |  |
| NSCLC | Bevacizumab,Carboplatin,Paclitaxel,Pembrolizumab | Yes |  |  |  |
| NSCLC | Bevacizumab,Carboplatin,Pemetrexed | Yes |  |  |  |
| NSCLC | Bevacizumab,Erlotinib | Yes |  |  |  |
| NSCLC | Bevacizumab,Osimertinib | Yes |  |  |  |
| NSCLC | Bevacizumab-Awwb | Yes |  |  |  |
| NSCLC | Bevacizumab-Awwb,Carboplatin,Osimertinib,Pemetrexed | Yes |  |  |  |
| NSCLC | Bevacizumab-Awwb,Carboplatin,Paclitaxel | Yes |  |  |  |
| NSCLC | Bevacizumab-Awwb,Carboplatin,Pemetrexed | Yes |  |  |  |
| NSCLC | Bevacizumab-Awwb,Osimertinib | Yes |  |  |  |
| NSCLC | Bevacizumab-Awwb,Pemetrexed | Yes |  |  |  |
| NSCLC | Bevacizumab-Bvzr,Carboplatin,Paclitaxel | Yes |  |  |  |
| NSCLC | Bevacizumab-Bvzr,Carboplatin,Pemetrexed | Yes |  |  |  |
| NSCLC | Bevacizumab-Bvzr,Erlotinib | Yes |  |  |  |
| NSCLC | Bicalutamide | Yes |  |  |  |
| NSCLC | Bicalutamide,Carboplatin,Leuprolide,Paclitaxel Protein-Bound,Pembrolizumab | Yes |  |  |  |
| NSCLC | Bicalutamide,Ipilimumab,Nivolumab | Yes |  |  |  |
| NSCLC | Bicalutamide,Leuprolide | Yes |  |  |  |
| NSCLC | Binimetinib,Carboplatin,Pembrolizumab,Pemetrexed | Yes |  |  |  |
| NSCLC | Binimetinib,Encorafenib | Yes |  |  |  |
| NSCLC | Bortezomib | Yes |  |  |  |
| NSCLC | Bortezomib,Carboplatin,Lenalidomide,Paclitaxel,Pembrolizumab | Yes |  |  |  |
| NSCLC | Bortezomib,Daratumumab | Yes |  |  |  |
| NSCLC | Brigatinib | Yes |  |  |  |
| NSCLC | Capecitabine | Yes |  |  |  |
| NSCLC | Capecitabine,Carboplatin,Osimertinib,Paclitaxel | Yes |  |  |  |
| NSCLC | Capmatinib | Yes |  |  |  |
| NSCLC | Carboplatin | Yes |  |  |  |
| NSCLC | Carboplatin,Clinical Study Drug,Paclitaxel,Pembrolizumab | Yes |  |  |  |
| NSCLC | Carboplatin,Clinical Study Drug,Pembrolizumab,Pemetrexed | Yes |  |  |  |
| NSCLC | Carboplatin,Clinical Study Drug,Pemetrexed | Yes |  |  |  |
| NSCLC | Carboplatin,Cyclophosphamide,Paclitaxel | Yes |  |  |  |
| NSCLC | Carboplatin,Cyclophosphamide,Pembrolizumab,Pemetrexed | Yes |  |  |  |
| NSCLC | Carboplatin,Docetaxel | Yes |  |  |  |
| NSCLC | Carboplatin,Docetaxel,Paclitaxel | Yes |  |  |  |
| NSCLC | Carboplatin,Docetaxel,Paclitaxel Protein-Bound | Yes |  |  |  |
| NSCLC | Carboplatin,Docetaxel,Paclitaxel,Pembrolizumab | Yes |  |  |  |
| NSCLC | Carboplatin,Docetaxel,Paclitaxel,Pemetrexed | Yes |  |  |  |
| NSCLC | Carboplatin,Docetaxel,Pembrolizumab | Yes |  |  |  |
| NSCLC | Carboplatin,Durvalumab,Gemcitabine | Yes |  |  |  |
| NSCLC | Carboplatin,Durvalumab,Pembrolizumab,Pemetrexed | Yes |  |  |  |
| NSCLC | Carboplatin,Etoposide | Yes |  |  |  |
| NSCLC | Carboplatin,Etoposide,Nivolumab | Yes |  |  |  |
| NSCLC | Carboplatin,Etoposide,Paclitaxel | Yes |  |  |  |
| NSCLC | Carboplatin,Etoposide,Pembrolizumab,Pemetrexed | Yes |  |  |  |
| NSCLC | Carboplatin,Etoposide,Pemetrexed | Yes |  |  |  |
| NSCLC | Carboplatin,Exemestane,Pembrolizumab,Pemetrexed | Yes |  |  |  |
| NSCLC | Carboplatin,Gemcitabine | Yes |  |  |  |
| NSCLC | Carboplatin,Gemcitabine,Paclitaxel | Yes |  |  |  |
| NSCLC | Carboplatin,Gemcitabine,Paclitaxel,Pembrolizumab | Yes |  |  |  |
| NSCLC | Carboplatin,Gemcitabine,Pembrolizumab | Yes |  |  |  |
| NSCLC | Carboplatin,Hydroxyurea,Pembrolizumab,Pemetrexed | Yes |  |  |  |
| NSCLC | Carboplatin,Ibrutinib,Pembrolizumab,Pemetrexed | Yes |  |  |  |
| NSCLC | Carboplatin,Ifosfamide,Mesna,Paclitaxel,Pembrolizumab,Pemetrexed | Yes |  |  |  |
| NSCLC | Carboplatin,Ipilimumab,Nivolumab,Paclitaxel | Yes |  |  |  |
| NSCLC | Carboplatin,Ipilimumab,Nivolumab,Paclitaxel Protein-Bound | Yes |  |  |  |
| NSCLC | Carboplatin,Ipilimumab,Nivolumab,Pemetrexed | Yes |  |  |  |
| NSCLC | Carboplatin,Lenalidomide,Pembrolizumab,Pemetrexed | Yes |  |  |  |
| NSCLC | Carboplatin,Letrozole,Pembrolizumab,Pemetrexed | Yes |  |  |  |
| NSCLC | Carboplatin,Leuprolide | Yes |  |  |  |
| NSCLC | Carboplatin,Leuprolide,Medroxyprogesterone,Pembrolizumab,Pemetrexed | Yes |  |  |  |
| NSCLC | Carboplatin,Leuprolide,Paclitaxel Protein-Bound | Yes |  |  |  |
| NSCLC | Carboplatin,Leuprolide,Pembrolizumab,Pemetrexed | Yes |  |  |  |
| NSCLC | Carboplatin,Nivolumab,Paclitaxel | Yes |  |  |  |
| NSCLC | Carboplatin,Nivolumab,Pembrolizumab,Pemetrexed | Yes |  |  |  |
| NSCLC | Carboplatin,Obinutuzumab,Paclitaxel,Pembrolizumab | Yes |  |  |  |
| NSCLC | Carboplatin,Osimertinib,Paclitaxel Protein-Bound,Pembrolizumab | Yes |  |  |  |
| NSCLC | Carboplatin,Osimertinib,Pembrolizumab,Pemetrexed | Yes |  |  |  |
| NSCLC | Carboplatin,Osimertinib,Pemetrexed | Yes |  |  |  |
| NSCLC | Carboplatin,Paclitaxel | Yes |  |  |  |
| NSCLC | Carboplatin,Paclitaxel Protein-Bound | Yes |  |  |  |
| NSCLC | Carboplatin,Paclitaxel Protein-Bound,Pembrolizumab | Yes |  |  |  |
| NSCLC | Carboplatin,Paclitaxel Protein-Bound,Pembrolizumab,Pemetrexed | Yes |  |  |  |
| NSCLC | Carboplatin,Paclitaxel,Pembrolizumab | Yes |  |  |  |
| NSCLC | Carboplatin,Paclitaxel,Pembrolizumab,Pemetrexed | Yes |  |  |  |
| NSCLC | Carboplatin,Paclitaxel,Pembrolizumab,Ruxolitinib | Yes |  |  |  |
| NSCLC | Carboplatin,Paclitaxel,Pemetrexed | Yes |  |  |  |
| NSCLC | Carboplatin,Pembrolizumab | Yes |  |  |  |
| NSCLC | Carboplatin,Pembrolizumab,Pemetrexed | Yes |  |  |  |
| NSCLC | Carboplatin,Pembrolizumab,Pemetrexed,Ruxolitinib | Yes |  |  |  |
| NSCLC | Carboplatin,Pembrolizumab,Pemetrexed,Thiotepa | Yes |  |  |  |
| NSCLC | Carboplatin,Pemetrexed | Yes |  |  |  |
| NSCLC | Carboplatin,Vinorelbine | Yes |  |  |  |
| NSCLC | Ceritinib | Yes |  |  |  |
| NSCLC | Cisplatin | Yes |  |  |  |
| NSCLC | Cisplatin,Docetaxel | Yes |  |  |  |
| NSCLC | Cisplatin,Docetaxel,Etoposide | Yes |  |  |  |
| NSCLC | Cisplatin,Docetaxel,Pembrolizumab | Yes |  |  |  |
| NSCLC | Cisplatin,Etoposide | Yes |  |  |  |
| NSCLC | Cisplatin,Gemcitabine | Yes |  |  |  |
| NSCLC | Cisplatin,Gemcitabine,Necitumumab | Yes |  |  |  |
| NSCLC | Cisplatin,Ipilimumab,Nivolumab,Pemetrexed | Yes |  |  |  |
| NSCLC | Cisplatin,Pembrolizumab,Pemetrexed | Yes |  |  |  |
| NSCLC | Cisplatin,Pemetrexed | Yes |  |  |  |
| NSCLC | Clinical Study Drug | Yes |  |  |  |
| NSCLC | Clinical Study Drug,Osimertinib | Yes |  |  |  |
| NSCLC | Copanlisib | Yes |  |  |  |
| NSCLC | Crizotinib | Yes |  |  |  |
| NSCLC | Crizotinib,Nivolumab | Yes |  |  |  |
| NSCLC | Cyclophosphamide | Yes |  |  |  |
| NSCLC | Cyclophosphamide,Doxorubicin,Etoposide,Rituximab-Abbs,Vincristine | Yes |  |  |  |
| NSCLC | Dabrafenib,Trametinib | Yes |  |  |  |
| NSCLC | Docetaxel | Yes |  |  |  |
| NSCLC | Docetaxel,Gemcitabine | Yes |  |  |  |
| NSCLC | Docetaxel,Ramucirumab | Yes |  |  |  |
| NSCLC | Durvalumab | Yes |  |  |  |
| NSCLC | Durvalumab,Pemetrexed | Yes |  |  |  |
| NSCLC | Durvalumab,Vinorelbine | Yes |  |  |  |
| NSCLC | Enfortumab Vedotin-Ejfv | Yes |  |  |  |
| NSCLC | Entrectinib | Yes |  |  |  |
| NSCLC | Erlotinib | Yes |  |  |  |
| NSCLC | Erlotinib,Ramucirumab | Yes |  |  |  |
| NSCLC | Etoposide | Yes |  |  |  |
| NSCLC | Everolimus | Yes |  |  |  |
| NSCLC | Exemestane | Yes |  |  |  |
| NSCLC | Exemestane,Ipilimumab,Nivolumab | Yes |  |  |  |
| NSCLC | Exemestane,Palbociclib | Yes |  |  |  |
| NSCLC | Fam-Trastuzumab Deruxtecan-Nxki | Yes |  |  |  |
| NSCLC | Fluorouracil,Irinotecan,Leucovorin,Oxaliplatin | Yes |  |  |  |
| NSCLC | Fluorouracil,Leucovorin,Oxaliplatin | Yes |  |  |  |
| NSCLC | Fulvestrant,Ipilimumab,Nivolumab,Ribociclib | Yes |  |  |  |
| NSCLC | Gefitinib | Yes |  |  |  |
| NSCLC | Gemcitabine | Yes |  |  |  |
| NSCLC | Gemcitabine,Paclitaxel | Yes |  |  |  |
| NSCLC | Gemcitabine,Paclitaxel Protein-Bound | Yes |  |  |  |
| NSCLC | Gemcitabine,Pembrolizumab | Yes |  |  |  |
| NSCLC | Gemcitabine,Vinorelbine | Yes |  |  |  |
| NSCLC | Hydroxyurea | Yes |  |  |  |
| NSCLC | Hydroxyurea,Pembrolizumab | Yes |  |  |  |
| NSCLC | Ibrutinib | Yes |  |  |  |
| NSCLC | Imatinib | Yes |  |  |  |
| NSCLC | Ipilimumab,Nivolumab | Yes |  |  |  |
| NSCLC | Ipilimumab,Nivolumab,Pemetrexed | Yes |  |  |  |
| NSCLC | Larotrectinib | Yes |  |  |  |
| NSCLC | Letrozole | Yes |  |  |  |
| NSCLC | Letrozole,Palbociclib | Yes |  |  |  |
| NSCLC | Letrozole,Pembrolizumab | Yes |  |  |  |
| NSCLC | Leuprolide | Yes |  |  |  |
| NSCLC | Methotrexate | Yes |  |  |  |
| NSCLC | Nivolumab | Yes |  |  |  |
| NSCLC | Olaparib | Yes |  |  |  |
| NSCLC | Osimertinib | Yes |  |  |  |
| NSCLC | Osimertinib,Pembrolizumab | Yes |  |  |  |
| NSCLC | Osimertinib,Rituximab | Yes |  |  |  |
| NSCLC | Paclitaxel | Yes |  |  |  |
| NSCLC | Paclitaxel Protein-Bound | Yes |  |  |  |
| NSCLC | Paclitaxel Protein-Bound,Pembrolizumab | Yes |  |  |  |
| NSCLC | Paclitaxel,Pembrolizumab | Yes |  |  |  |
| NSCLC | Palbociclib | Yes |  |  |  |
| NSCLC | Palbociclib,Pembrolizumab | Yes |  |  |  |
| NSCLC | Pazopanib | Yes |  |  |  |
| NSCLC | Pembrolizumab | Yes |  |  |  |
| NSCLC | Pembrolizumab,Pemetrexed | Yes |  |  |  |
| NSCLC | Pembrolizumab,Ruxolitinib | Yes |  |  |  |
| NSCLC | Pembrolizumab,Vinorelbine | Yes |  |  |  |
| NSCLC | Pemetrexed | Yes |  |  |  |
| NSCLC | Rituximab | Yes |  |  |  |
| NSCLC | Rituximab-Pvvr | Yes |  |  |  |
| NSCLC | Ruxolitinib | Yes |  |  |  |
| NSCLC | Selpercatinib | Yes |  |  |  |
| NSCLC | Tamoxifen | Yes |  |  |  |
| NSCLC | Temozolomide | Yes |  |  |  |
| NSCLC | Thiotepa | Yes |  |  |  |
| NSCLC | Trastuzumab | Yes |  |  |  |
| NSCLC | Vemurafenib | Yes |  |  |  |
| NSCLC | Vinorelbine | Yes |  |  |  |
| UCC | Capecitabine | Yes |  |  |  |
| UCC | Tamoxifen | Yes |  |  |  |
| UCC | Acalabrutinib | Yes |  |  |  |
| UCC | Ado-Trastuzumab Emtansine | Yes |  |  |  |
| UCC | Anastrozole | Yes |  |  |  |
| UCC | Axitinib,Imatinib | Yes |  |  |  |
| UCC | Bcg Vaccine | Yes |  |  |  |
| UCC | Fluorouracil,Mitomycin | Yes |  |  |  |
| UCC | Hydroxyurea | Yes |  |  |  |
| UCC | Ibrutinib | Yes |  |  |  |
| UCC | Letrozole | Yes |  |  |  |
| UCC | Leuprolide | Yes |  |  |  |
| UCC | Mitomycin | Yes |  |  |  |
| UCC | Rituximab | Yes |  |  |  |
| UCC | Triptorelin | Yes |  |  |  |
| UCC | Abiraterone,Carboplatin,Gemcitabine | Yes |  |  |  |
| UCC | Capecitabine,Carboplatin,Gemcitabine,Mitomycin | Yes |  |  |  |
| UCC | Carboplatin,Enfortumab Vedotin-Ejfv,Gemcitabine | Yes |  |  |  |
| UCC | Anastrozole,Pembrolizumab | Yes |  |  |  |
| UCC | Atezolizumab | Yes |  |  |  |
| UCC | Atezolizumab,Carboplatin,Etoposide | Yes |  |  |  |
| UCC | Atezolizumab,Carboplatin,Gemcitabine | Yes |  |  |  |
| UCC | Atezolizumab,Cisplatin,Gemcitabine | Yes |  |  |  |
| UCC | Atezolizumab,Gemcitabine,Paclitaxel | Yes |  |  |  |
| UCC | Atezolizumab,Pembrolizumab | Yes |  |  |  |
| UCC | Avelumab | Yes |  |  |  |
| UCC | Bevacizumab,Gemcitabine | Yes |  |  |  |
| UCC | Carboplatin,Etoposide | Yes |  |  |  |
| UCC | Carboplatin,Gemcitabine | Yes |  |  |  |
| UCC | Carboplatin,Gemcitabine,Paclitaxel Protein-Bound | Yes |  |  |  |
| UCC | Carboplatin,Gemcitabine,Paclitaxel,Pembrolizumab | Yes |  |  |  |
| UCC | Carboplatin,Gemcitabine,Pembrolizumab | Yes |  |  |  |
| UCC | Carboplatin,Paclitaxel | Yes |  |  |  |
| UCC | Carboplatin,Paclitaxel,Paclitaxel Protein-Bound | Yes |  |  |  |
| UCC | Carboplatin,Pralatrexate | Yes |  |  |  |
| UCC | Cisplatin | Yes |  |  |  |
| UCC | Cisplatin,Etoposide | Yes |  |  |  |
| UCC | Cisplatin,Fluorouracil | Yes |  |  |  |
| UCC | Cisplatin,Fluorouracil,Gemcitabine | Yes |  |  |  |
| UCC | Cisplatin,Gemcitabine | Yes |  |  |  |
| UCC | Cisplatin,Gemcitabine,Ifosfamide | Yes |  |  |  |
| UCC | Cisplatin,Gemcitabine,Paclitaxel | Yes |  |  |  |
| UCC | Cisplatin,Gemcitabine,Pembrolizumab | Yes |  |  |  |
| UCC | Cisplatin,Gemcitabine,Sunitinib | Yes |  |  |  |
| UCC | Cisplatin,Paclitaxel | Yes |  |  |  |
| UCC | Docetaxel,Gemcitabine | Yes |  |  |  |
| UCC | Doxorubicin,Gemcitabine,Paclitaxel | Yes |  |  |  |
| UCC | Durvalumab | Yes |  |  |  |
| UCC | Enfortumab Vedotin-Ejfv | Yes |  |  |  |
| UCC | Enfortumab Vedotin-Ejfv,Pembrolizumab | Yes |  |  |  |
| UCC | Erdafitinib | Yes |  |  |  |
| UCC | Gemcitabine | Yes |  |  |  |
| UCC | Gemcitabine,Oxaliplatin | Yes |  |  |  |
| UCC | Gemcitabine,Paclitaxel | Yes |  |  |  |
| UCC | Gemcitabine,Pembrolizumab | Yes |  |  |  |
| UCC | Hydroxyurea,Pembrolizumab | Yes |  |  |  |
| UCC | Imatinib,Pembrolizumab | Yes |  |  |  |
| UCC | MVAC | Yes |  |  |  |
| UCC | MVAC,Pembrolizumab | Yes |  |  |  |
| UCC | Nivolumab | Yes |  |  |  |
| UCC | Paclitaxel | Yes |  |  |  |
| UCC | Paclitaxel Protein-Bound | Yes |  |  |  |
| UCC | Paclitaxel,Tamoxifen | Yes |  |  |  |
| UCC | Pembrolizumab | Yes |  |  |  |
| UCC | Pembrolizumab,Thalidomide | Yes |  |  |  |
| UCC | Clinical Study Drug,Pembrolizumab | Yes |  |  |  |
| UCC | Carboplatin,Clinical Study Drug | Yes |  |  |  |
| UCC | Carboplatin,Clinical Study Drug,Gemcitabine | Yes |  |  |  |
| UCC | Cisplatin,Clinical Study Drug,Enfortumab Vedotin-Ejfv | Yes |  |  |  |
| UCC | Cisplatin,Clinical Study Drug,Gemcitabine | Yes |  |  |  |
| UCC | Clinical Study Drug | Yes |  |  |  |
| Prostate | Lenalidomide,Medroxyprogesterone,Rituximab | Yes |  |  |  |
| Prostate | Rituximab | Yes |  |  |  |
| Prostate | Exemestane | Yes |  |  |  |
| Prostate | Ibrutinib | Yes |  |  |  |
| Prostate | Irinotecan | Yes |  |  |  |
| Prostate | Abiraterone,Cemiplimab | Yes |  |  |  |
| Prostate | Abiraterone,Enzalutamide | Yes |  |  |  |
| Prostate | Abiraterone,Enzalutamide,Paclitaxel | Yes |  |  |  |
| Prostate | Abiraterone,Radium-223 | Yes |  |  |  |
| Prostate | Apalutamide,Docetaxel,Enzalutamide | Yes |  |  |  |
| Prostate | Apalutamide,Docetaxel,Paclitaxel | Yes |  |  |  |
| Prostate | Apalutamide,Enzalutamide | Yes |  |  |  |
| Prostate | Cabazitaxel,Enzalutamide | Yes |  |  |  |
| Prostate | Carboplatin,Docetaxel | Yes |  |  |  |
| Prostate | Carboplatin,Docetaxel,Olaparib | Yes |  |  |  |
| Prostate | Darolutamide | Yes |  |  |  |
| Prostate | Darolutamide,Sipuleucel-T | Yes |  |  |  |
| Prostate | Diethylstilbestrol | Yes |  |  |  |
| Prostate | Enzalutamide,Paclitaxel | Yes |  |  |  |
| Prostate | Ketoconazole | Yes |  |  |  |
| Prostate | Niraparib,Rucaparib | Yes |  |  |  |
| Prostate | Olaparib | Yes |  |  |  |
| Prostate | Pembrolizumab | Yes |  |  |  |
| Prostate | Abiraterone | Yes |  |  |  |
| Prostate | Abiraterone,Docetaxel | Yes |  |  |  |
| Prostate | Abiraterone,Rituximab | Yes |  |  |  |
| Prostate | Abiraterone,Sipuleucel-T | Yes |  |  |  |
| Prostate | Apalutamide | Yes |  |  |  |
| Prostate | Cabazitaxel | Yes |  |  |  |
| Prostate | Carboplatin,Enzalutamide,Etoposide | Yes |  |  |  |
| Prostate | Carboplatin,Etoposide | Yes |  |  |  |
| Prostate | Docetaxel | Yes |  |  |  |
| Prostate | Docetaxel,Enzalutamide | Yes |  |  |  |
| Prostate | Docetaxel,Nivolumab | Yes |  |  |  |
| Prostate | Enzalutamide | Yes |  |  |  |
| Prostate | Enzalutamide,Radium-223 | Yes |  |  |  |
| Prostate | Radium-223 | Yes |  |  |  |
| Prostate | Sipuleucel-T | Yes |  |  |  |
| Prostate | Abiraterone,Clinical Study Drug | Yes |  |  |  |
| Prostate | Clinical Study Drug | Yes |  |  |  |
| RCC | Aldesleukin | No | Yes | No | No |
| RCC | Azacitidine | No | No |  |  |
| RCC | Bevacizumab-Awwb,Cisplatin,Pemetrexed | No | No |  |  |
| RCC | Cabozantinib,Nivolumab | No | Yes | No | No |
| RCC | Cyclophosphamide,Doxorubicin,Rituximab-Pvvr,Vincristine | No | No |  |  |
| RCC | Imatinib | No | No |  |  |
| RCC | Lenalidomide | No | No |  |  |
| RCC | Lenvatinib,Pembrolizumab | No | Yes | No | No |
| RCC | Peginterferon Alfa-2B | No | Yes | No | No |
| RCC | Anastrozole | No | No |  |  |
| RCC | Avelumab | No | Yes | No | No |
| RCC | Avelumab,Axitinib | No | Yes | No | No |
| RCC | Avelumab,Axitinib,Ipilimumab,Nivolumab | No | Yes | No | No |
| RCC | Axitinib | No | Yes | No | No |
| RCC | Axitinib,Ipilimumab,Nivolumab | No | Yes | No | No |
| RCC | Axitinib,Ipilimumab,Nivolumab,Pembrolizumab | No | Yes | No | No |
| RCC | Axitinib,Leuprolide,Nivolumab | No | Yes | No | No |
| RCC | Axitinib,Nivolumab | No | Yes | No | No |
| RCC | Axitinib,Nivolumab,Pembrolizumab | No | Yes | No | No |
| RCC | Axitinib,Pembrolizumab | No | Yes | No | No |
| RCC | Bevacizumab,Erlotinib | No | No |  |  |
| RCC | Bevacizumab,Nivolumab | No | Yes | No | No |
| RCC | Bicalutamide,Leuprolide,Sunitinib | No | Yes | No | No |
| RCC | Cabazitaxel,Cabozantinib | No | Yes | No | No |
| RCC | Cabozantinib | No | Yes | No | No |
| RCC | Cabozantinib,Ipilimumab,Nivolumab | No | Yes | No | No |
| RCC | Capecitabine | No | No |  |  |
| RCC | Carboplatin,Etoposide | No | No |  |  |
| RCC | Carboplatin,Paclitaxel | No | No |  |  |
| RCC | Carboplatin,Pemetrexed | No | No |  |  |
| RCC | Cisplatin,Doxorubicin,Methotrexate,Vinblastine | No | No |  |  |
| RCC | Cisplatin,Gemcitabine | No | No |  |  |
| RCC | Clinical Study Drug | Yes |  |  |  |
| RCC | Clinical Study Drug,Gemcitabine | Yes |  |  |  |
| RCC | Clinical Study Drug,Ipilimumab,Nivolumab | Yes |  |  |  |
| RCC | Clinical Study Drug,Pembrolizumab | Yes |  |  |  |
| RCC | Cyclophosphamide | No | No |  |  |
| RCC | Dasatinib | No | No |  |  |
| RCC | Durvalumab | No | No |  |  |
| RCC | Everolimus,Lenvatinib | No | Yes | No | No |
| RCC | Ipilimumab,Leuprolide,Nivolumab | No | Yes | No | No |
| RCC | Ipilimumab,Medroxyprogesterone,Nivolumab | No | Yes | No | No |
| RCC | Ipilimumab,Nivolumab | No | Yes | No | No |
| RCC | Ipilimumab,Nivolumab,Pazopanib | No | Yes | No | No |
| RCC | Ipilimumab,Nivolumab,Pembrolizumab | No | Yes | No | No |
| RCC | Ipilimumab,Nivolumab,Sunitinib | No | Yes | No | No |
| RCC | Lenvatinib | No | Yes | No | No |
| RCC | Nivolumab | No | Yes | No | No |
| RCC | Nivolumab,Pazopanib | No | Yes | No | No |
| RCC | Nivolumab,Pembrolizumab | No | Yes | No | No |
| RCC | Paclitaxel,Sunitinib | No | Yes | No | No |
| RCC | Pazopanib | No | Yes | No | No |
| RCC | Pembrolizumab | No | Yes | No | No |
| RCC | Rituximab | No | No |  |  |
| RCC | Sorafenib | No | Yes | No | No |
| RCC | Sunitinib | No | Yes | No | No |
| RCC | Temsirolimus | No | Yes | No | No |
| CRC | Bevacizumab-Bvzr | No | Yes | No | No |
| CRC | Binimetinib,Encorafenib,Panitumumab | No | Yes | No | No |
| CRC | Capecitabine,Ipilimumab,Nivolumab | No | Yes | No | No |
| CRC | CAPEOX,Cetuximab,Panitumumab | No | Yes | No | No |
| CRC | CAPEOX,Ipilimumab,Nivolumab,Pembrolizumab | No | Yes | No | No |
| CRC | Cisplatin,Vinblastine,Vinorelbine | No | No |  |  |
| CRC | Cyclophosphamide,Doxorubicin,Rituximab/Hyaluronidase,Vincristine | No | No |  |  |
| CRC | Cyclophosphamide,Rituximab/Hyaluronidase | No | No |  |  |
| CRC | Degarelix,Enzalutamide | No | No |  |  |
| CRC | Encorafenib,Cetuximab | No | Yes | No | No |
| CRC | Encorafenib,Oxaliplatin,Cetuximab | No | Yes | No | No |
| CRC | Encorafenib,Panitumumab | No | Yes | No | No |
| CRC | FOLFIRI,Bicalutamide,Leuprolide | No | Yes | No | No |
| CRC | FOLFIRI,Clinical Study Drug | Yes |  |  |  |
| CRC | FOLFOX,Bevacizumab-Awwb,Cetuximab | No | Yes | No | No |
| CRC | FOLFOX,Tamoxifen,Bevacizumab-Bvzr | No | Yes | No | No |
| CRC | FOLFOXIRI,Bevacizumab-Awwb,Cetuximab | No | Yes | No | No |
| CRC | FOLFOXIRI,Bevacizumab-Bvzr | No | Yes | No | No |
| CRC | Leuprolide | No | No |  |  |
| CRC | Palbociclib | No | No |  |  |
| CRC | Sorafenib | No | No |  |  |
| CRC | Trifluridine/Tipiracil,Panitumumab | No | Yes | No | No |
| CRC | Abiraterone | No | No |  |  |
| CRC | Acalabrutinib | No | Yes | No | No |
| CRC | Azacitidine,Cetuximab | No | Yes | No | No |
| CRC | Bevacizumab | No | Yes | No | No |
| CRC | Bevacizumab-Awwb | No | Yes | No | No |
| CRC | Binimetinib,Dabrafenib,Trametinib,Cetuximab | No | Yes | No | No |
| CRC | Binimetinib,Encorafenib,Cetuximab | No | Yes | No | No |
| CRC | Binimetinib,Panitumumab | No | Yes | No | No |
| CRC | Capecitabine | No | Yes | No | No |
| CRC | Capecitabine,Bevacizumab | No | Yes | No | No |
| CRC | Capecitabine,Bevacizumab-Awwb | No | Yes | No | No |
| CRC | Capecitabine,Bevacizumab-Bvzr | No | Yes | No | No |
| CRC | Capecitabine,Irinotecan | No | Yes | No | No |
| CRC | Capecitabine,Irinotecan,Bevacizumab | No | Yes | No | No |
| CRC | Capecitabine,Irinotecan,Bevacizumab-Awwb | No | Yes | No | Yes |
| CRC | Capecitabine,Methotrexate | No | Yes | No | No |
| CRC | Capecitabine,Nivolumab | No | Yes | No | No |
| CRC | Capecitabine,Pembrolizumab | No | Yes | No | Yes |
| CRC | CAPEOX | No | Yes | No | No |
| CRC | CAPEOX,Bevacizumab | No | Yes | No | No |
| CRC | CAPEOX,Bevacizumab-Awwb | No | Yes | No | No |
| CRC | CAPEOX,Bevacizumab-Bvzr | No | Yes | No | No |
| CRC | CAPEOX,Cetuximab | No | Yes | No | No |
| CRC | CAPEOX,Imatinib,Bevacizumab | No | Yes | No | No |
| CRC | CAPEOX,Panitumumab | No | Yes | No | No |
| CRC | CAPEOX,Pembrolizumab,Bevacizumab-Awwb | No | Yes | No | Yes |
| CRC | Carboplatin,Paclitaxel | No | No |  |  |
| CRC | Carboplatin,Paclitaxel,Bevacizumab | No | No |  |  |
| CRC | Carboplatin,Paclitaxel,Cetuximab | No | Yes | No | Yes |
| CRC | Cetuximab | No | Yes | No | No |
| CRC | Cisplatin | No | No |  |  |
| CRC | Cisplatin,Gemcitabine | No | No |  |  |
| CRC | Clinical Study Drug | Yes |  |  |  |
| CRC | Degarelix,Leuprolide | No | No |  |  |
| CRC | Exemestane | No | No |  |  |
| CRC | Fluorouracil | No | Yes | No | No |
| CRC | Fluorouracil,Bevacizumab | No | Yes | No | No |
| CRC | Fluorouracil,Bleomycin,Cisplatin,Clinical Study Drug,Etoposide | Yes |  |  |  |
| CRC | Fluorouracil,Cetuximab | No | Yes | No | No |
| CRC | Fluorouracil,Cisplatin | No | Yes | No | Yes |
| CRC | Fluorouracil,Leucovorin | No | Yes | No | No |
| CRC | Fluorouracil,Leucovorin,Bevacizumab | No | Yes | No | No |
| CRC | Fluorouracil,Leucovorin,Bevacizumab-Awwb | No | Yes | No | No |
| CRC | Fluorouracil,Leucovorin,Bevacizumab,Panitumumab | No | Yes | No | No |
| CRC | Fluorouracil,Leucovorin,Leuprolide | No | Yes | No | Yes |
| CRC | Fluorouracil,Leucovorin,Panitumumab | No | Yes | No | Yes |
| CRC | Fluorouracil,Leucovorin,Tamoxifen | No | Yes | No | No |
| CRC | FOLFIRI | No | Yes | No | No |
| CRC | FOLFIRI,Bevacizumab | No | Yes | No | No |
| CRC | FOLFIRI,Bevacizumab-Awwb | No | Yes | No | No |
| CRC | FOLFIRI,Bevacizumab-Bvzr | No | Yes | No | No |
| CRC | FOLFIRI,Bevacizumab,Cetuximab | No | Yes | No | Yes |
| CRC | FOLFIRI,Bevacizumab,Cetuximab,Panitumumab | No | Yes | No | No |
| CRC | FOLFIRI,Bevacizumab,Panitumumab | No | Yes | No | No |
| CRC | FOLFIRI,Cetuximab | No | Yes | No | No |
| CRC | FOLFIRI,Enzalutamide,Cetuximab | No | Yes | No | No |
| CRC | FOLFIRI,Leuprolide,Bevacizumab | No | Yes | No | No |
| CRC | FOLFIRI,Olaparib,Bevacizumab-Awwb | No | Yes | No | No |
| CRC | FOLFIRI,Panitumumab | No | Yes | No | No |
| CRC | FOLFIRI,Pembrolizumab | No | Yes | No | Yes |
| CRC | FOLFIRI,Ziv-Aflibercept | No | Yes | No | No |
| CRC | FOLFOX | No | Yes | No | No |
| CRC | FOLFOX,Abiraterone,Leuprolide,Bevacizumab | No | Yes | No | No |
| CRC | FOLFOX,Bevacizumab | No | Yes | No | No |
| CRC | FOLFOX,Bevacizumab-Awwb | No | Yes | No | No |
| CRC | FOLFOX,Bevacizumab-Awwb,Panitumumab | No | Yes | No | No |
| CRC | FOLFOX,Bevacizumab-Bvzr | No | Yes | No | No |
| CRC | FOLFOX,Bevacizumab,Cetuximab | No | Yes | No | No |
| CRC | FOLFOX,Bevacizumab,Cetuximab,Panitumumab | No | Yes | No | No |
| CRC | FOLFOX,Bevacizumab,Panitumumab | No | Yes | No | No |
| CRC | FOLFOX,Cetuximab | No | Yes | No | No |
| CRC | FOLFOX,Cetuximab,Panitumumab | No | Yes | No | No |
| CRC | FOLFOX,Cisplatin,Gemcitabine,Bevacizumab | No | Yes | No | No |
| CRC | FOLFOX,Clinical Study Drug | Yes |  |  |  |
| CRC | FOLFOX,Clinical Study Drug,Bevacizumab | Yes |  |  |  |
| CRC | FOLFOX,Clinical Study Drug,Bevacizumab-Awwb | Yes |  |  |  |
| CRC | FOLFOX,Daratumumab,Pomalidomide | No | Yes | No | No |
| CRC | FOLFOX,Hydroxyurea | No | Yes | No | No |
| CRC | FOLFOX,Letrozole | No | Yes | No | No |
| CRC | FOLFOX,Leuprolide,Bevacizumab | No | Yes | No | No |
| CRC | FOLFOX,Panitumumab | No | Yes | No | No |
| CRC | FOLFOXIRI | No | Yes | No | No |
| CRC | FOLFOXIRI,Bevacizumab | No | Yes | No | No |
| CRC | FOLFOXIRI,Bevacizumab-Awwb | No | Yes | No | No |
| CRC | FOLFOXIRI,Bevacizumab-Awwb,Panitumumab | No | Yes | No | No |
| CRC | FOLFOXIRI,Cetuximab | No | Yes | No | No |
| CRC | FOLFOXIRI,Panitumumab | No | Yes | No | No |
| CRC | Fulvestrant,Palbociclib | No | No |  |  |
| CRC | Gemcitabine,Paclitaxel Protein-Bound | No | Yes | No | Yes |
| CRC | Goserelin,Trastuzumab-Anns | No | No |  |  |
| CRC | Hydroxyurea | No | No |  |  |
| CRC | Ibrutinib | No | No |  |  |
| CRC | Ibrutinib,Pembrolizumab | No | Yes | Yes |  |
| CRC | Imatinib | No | No |  |  |
| CRC | Ipilimumab,Nivolumab | No | Yes | No | No |
| CRC | Irinotecan | No | Yes | No | Yes |
| CRC | Irinotecan,Bevacizumab | No | Yes | No | No |
| CRC | Irinotecan,Bevacizumab-Awwb | No | Yes | No | Yes |
| CRC | Irinotecan,Cetuximab | No | Yes | No | No |
| CRC | Irinotecan,Oxaliplatin,Bevacizumab | No | Yes | No | Yes |
| CRC | Irinotecan,Panitumumab | No | Yes | No | No |
| CRC | Irinotecan,Ramucirumab | No | Yes | No | Yes |
| CRC | Lenvatinib | No | No |  |  |
| CRC | Letrozole | No | No |  |  |
| CRC | Medroxyprogesterone | No | No |  |  |
| CRC | Methotrexate | No | No |  |  |
| CRC | Nivolumab | No | Yes | No | No |
| CRC | Nivolumab,Pembrolizumab | No | Yes | Yes |  |
| CRC | Oxaliplatin | No | Yes | No | No |
| CRC | Oxaliplatin,Bevacizumab | No | Yes | No | No |
| CRC | Oxaliplatin,Bevacizumab-Awwb | No | Yes | No | No |
| CRC | Panitumumab | No | Yes | No | No |
| CRC | Pembrolizumab | No | Yes | Yes |  |
| CRC | Regorafenib | No | Yes | No | No |
| CRC | Tamoxifen | No | No |  |  |
| CRC | Temozolomide,Bevacizumab | No | No |  |  |
| CRC | Tretinoin | No | No |  |  |
| CRC | Trifluridine/Tipiracil | No | No |  |  |
| CRC | Venetoclax | No | No |  |  |
| Pancreatic | Bendamustine,Rituximab | No | No |  |  |
| Pancreatic | Capecitabine,Irinotecan Liposomal,Leucovorin | No | Yes | No | No |
| Pancreatic | Carboplatin,Fluorouracil,Irinotecan,Leucovorin,Oxaliplatin,Pembrolizumab,Pemetrexed | No | Yes | No | No |
| Pancreatic | Niraparib | No | No |  |  |
| Pancreatic | Afatinib | No | No |  |  |
| Pancreatic | Alpelisib | No | No |  |  |
| Pancreatic | Anastrozole | No | No |  |  |
| Pancreatic | Anastrozole,Capecitabine | No | Yes | No | No |
| Pancreatic | Anastrozole,Carboplatin,Fluorouracil,Irinotecan,Leucovorin,Oxaliplatin,Pembrolizumab,Pemetrexed | No | Yes | No | No |
| Pancreatic | Anastrozole,Fluorouracil,Irinotecan,Leucovorin,Oxaliplatin | No | Yes | No | No |
| Pancreatic | Anastrozole,Gemcitabine,Paclitaxel Protein-Bound | No | Yes | No | No |
| Pancreatic | Atezolizumab | No | No |  |  |
| Pancreatic | Axitinib,Gemcitabine | No | Yes | No | No |
| Pancreatic | Bevacizumab-Awwb,Capecitabine,Irinotecan | No | Yes | No | No |
| Pancreatic | Bevacizumab-Awwb,Fluorouracil,Leucovorin,Oxaliplatin | No | Yes | No | No |
| Pancreatic | Bevacizumab,Capecitabine,Irinotecan,Leucovorin,Oxaliplatin | No | Yes | No | No |
| Pancreatic | Bevacizumab,Carboplatin,Pemetrexed | No | No |  |  |
| Pancreatic | Bevacizumab,Fluorouracil,Leucovorin,Oxaliplatin | No | Yes | No | No |
| Pancreatic | Bevacizumab,Gemcitabine,Paclitaxel Protein-Bound | No | Yes | No | No |
| Pancreatic | Bevacizumab,Paclitaxel Protein-Bound,Pembrolizumab | No | Yes | No | No |
| Pancreatic | Bicalutamide | No | No |  |  |
| Pancreatic | Bicalutamide,Gemcitabine | No | Yes | No | No |
| Pancreatic | Bortezomib,Fluorouracil,Irinotecan,Leucovorin,Oxaliplatin | No | Yes | No | No |
| Pancreatic | Capecitabine | No | Yes | No | No |
| Pancreatic | Capecitabine,Cisplatin,Docetaxel,Gemcitabine | No | Yes | No | No |
| Pancreatic | Capecitabine,Cisplatin,Gemcitabine | No | Yes | No | No |
| Pancreatic | Capecitabine,Clinical Study Drug | Yes |  |  |  |
| Pancreatic | Capecitabine,Clinical Study Drug,Leucovorin,Oxaliplatin | Yes |  |  |  |
| Pancreatic | Capecitabine,Docetaxel,Gemcitabine | No | Yes | No | No |
| Pancreatic | Capecitabine,Docetaxel,Gemcitabine,Paclitaxel Protein-Bound | No | Yes | No | No |
| Pancreatic | Capecitabine,Erlotinib | No | Yes | No | No |
| Pancreatic | Capecitabine,Gemcitabine | No | Yes | No | No |
| Pancreatic | Capecitabine,Gemcitabine,Irinotecan,Leucovorin,Oxaliplatin,Paclitaxel Protein-Bound | No | Yes | No | No |
| Pancreatic | Capecitabine,Gemcitabine,Mitomycin | No | Yes | No | No |
| Pancreatic | Capecitabine,Gemcitabine,Paclitaxel Protein-Bound | No | Yes | No | No |
| Pancreatic | Capecitabine,Gemcitabine,Paclitaxel Protein-Bound,Ruxolitinib | No | Yes | No | No |
| Pancreatic | Capecitabine,Gemcitabine,Paclitaxel Protein-Bound,Temozolomide | No | Yes | No | No |
| Pancreatic | Capecitabine,Irinotecan Liposomal | No | Yes | No | No |
| Pancreatic | Capecitabine,Irinotecan,Leucovorin,Oxaliplatin | No | Yes | No | No |
| Pancreatic | Capecitabine,Irinotecan,Oxaliplatin | No | Yes | No | No |
| Pancreatic | Capecitabine,Leucovorin | No | Yes | No | No |
| Pancreatic | Capecitabine,Mitomycin | No | Yes | No | No |
| Pancreatic | Capecitabine,Temozolomide | No | Yes | No | No |
| Pancreatic | CAPEOX | No | Yes | No | No |
| Pancreatic | Carboplatin | No | No |  |  |
| Pancreatic | Carboplatin,Etoposide | No | No |  |  |
| Pancreatic | Carboplatin,Gemcitabine | No | Yes | No | No |
| Pancreatic | Carboplatin,Gemcitabine,Paclitaxel Protein-Bound | No | Yes | No | No |
| Pancreatic | Carboplatin,Gemcitabine,Paclitaxel Protein-Bound,Pembrolizumab,Pemetrexed | No | Yes | No | No |
| Pancreatic | Carboplatin,Gemcitabine,Paclitaxel,Paclitaxel Protein-Bound | No | Yes | No | No |
| Pancreatic | Carboplatin,Irinotecan | No | Yes | No | No |
| Pancreatic | Carboplatin,Paclitaxel | No | Yes | No | No |
| Pancreatic | Carboplatin,Paclitaxel Protein-Bound | No | Yes | No | No |
| Pancreatic | Carboplatin,Pembrolizumab,Pemetrexed | No | No |  |  |
| Pancreatic | Carboplatin,Pemetrexed | No | No |  |  |
| Pancreatic | Cemiplimab,Irinotecan | No | Yes | No | No |
| Pancreatic | Chlorambucil,Fluorouracil,Leucovorin,Obinutuzumab,Oxaliplatin | No | Yes | No | No |
| Pancreatic | Cisplatin | No | Yes | No | No |
| Pancreatic | Cisplatin,Docetaxel,Fluorouracil,Leucovorin | No | Yes | No | No |
| Pancreatic | Cisplatin,Erlotinib,Gemcitabine,Paclitaxel Protein-Bound | No | Yes | No | No |
| Pancreatic | Cisplatin,Fluorouracil,Irinotecan,Leucovorin,Oxaliplatin | No | Yes | No | No |
| Pancreatic | Cisplatin,Fluorouracil,Irinotecan,Leucovorin,Paclitaxel Protein-Bound | No | Yes | No | No |
| Pancreatic | Cisplatin,Fluorouracil,Trastuzumab | No | Yes | No | No |
| Pancreatic | Cisplatin,Gemcitabine | No | Yes | No | No |
| Pancreatic | Cisplatin,Gemcitabine,Nivolumab,Paclitaxel Protein-Bound | No | Yes | No | No |
| Pancreatic | Cisplatin,Gemcitabine,Olaparib | No | Yes | No | No |
| Pancreatic | Cisplatin,Gemcitabine,Oxaliplatin | No | Yes | No | No |
| Pancreatic | Cisplatin,Gemcitabine,Paclitaxel | No | Yes | No | No |
| Pancreatic | Cisplatin,Gemcitabine,Paclitaxel Protein-Bound | No | Yes | No | No |
| Pancreatic | Cisplatin,Interferon Alfa-2B,Irinotecan,Paclitaxel Protein-Bound | No | Yes | No | No |
| Pancreatic | Cisplatin,Irinotecan,Mitomycin | No | Yes | No | No |
| Pancreatic | Cisplatin,Vinorelbine | No | Yes | No | No |
| Pancreatic | Clinical Study Drug | Yes |  |  |  |
| Pancreatic | Clinical Study Drug,Fluorouracil,Irinotecan,Leucovorin | Yes |  |  |  |
| Pancreatic | Clinical Study Drug,Fluorouracil,Irinotecan,Leucovorin,Oxaliplatin | Yes |  |  |  |
| Pancreatic | Clinical Study Drug,Gemcitabine | Yes |  |  |  |
| Pancreatic | Clinical Study Drug,Gemcitabine,Paclitaxel Protein-Bound | Yes |  |  |  |
| Pancreatic | Clinical Study Drug,Irinotecan,Leucovorin,Oxaliplatin | Yes |  |  |  |
| Pancreatic | Clinical Study Drug,Paclitaxel Protein-Bound | Yes |  |  |  |
| Pancreatic | Clinical Study Drug,Trifluridine/Tipiracil | Yes |  |  |  |
| Pancreatic | Crizotinib,Gemcitabine,Paclitaxel Protein-Bound | No | Yes | No | No |
| Pancreatic | Cyclophosphamide | No | No |  |  |
| Pancreatic | Dacarbazine,Docetaxel,Fluorouracil,Gemcitabine,Irinotecan,Leucovorin,Oxaliplatin | No | Yes | No | No |
| Pancreatic | Dacarbazine,Doxorubicin,Fluorouracil,Irinotecan,Leucovorin,Oxaliplatin,Vinblastine | No | Yes | No | No |
| Pancreatic | Dasatinib | No | No |  |  |
| Pancreatic | Dasatinib,Fluorouracil,Irinotecan,Leucovorin,Oxaliplatin | No | Yes | No | No |
| Pancreatic | Docetaxel,Fluorouracil,Gemcitabine,Irinotecan,Leucovorin,Oxaliplatin | No | Yes | No | No |
| Pancreatic | Docetaxel,Fluorouracil,Irinotecan,Leucovorin,Oxaliplatin | No | Yes | No | No |
| Pancreatic | Docetaxel,Gemcitabine | No | Yes | No | No |
| Pancreatic | Enzalutamide | No | No |  |  |
| Pancreatic | Erlotinib | No | No |  |  |
| Pancreatic | Erlotinib,Fluorouracil,Gemcitabine,Paclitaxel Protein-Bound | No | Yes | No | No |
| Pancreatic | Erlotinib,Gemcitabine | No | Yes | No | No |
| Pancreatic | Erlotinib,Gemcitabine,Paclitaxel Protein-Bound | No | Yes | No | No |
| Pancreatic | Everolimus,Fluorouracil,Irinotecan Liposomal,Leucovorin | No | Yes | No | No |
| Pancreatic | Exemestane,Palbociclib | No | No |  |  |
| Pancreatic | Fluorouracil | No | Yes | No | No |
| Pancreatic | Fluorouracil,Gemcitabine | No | Yes | No | No |
| Pancreatic | Fluorouracil,Gemcitabine,Irinotecan Liposomal,Leucovorin | No | Yes | No | No |
| Pancreatic | Fluorouracil,Gemcitabine,Irinotecan Liposomal,Leucovorin,Paclitaxel Protein-Bound | No | Yes | No | No |
| Pancreatic | Fluorouracil,Gemcitabine,Irinotecan,Leucovorin | No | Yes | No | No |
| Pancreatic | Fluorouracil,Gemcitabine,Irinotecan,Leucovorin,Oxaliplatin | No | Yes | No | No |
| Pancreatic | Fluorouracil,Gemcitabine,Irinotecan,Leucovorin,Oxaliplatin,Paclitaxel Protein-Bound | No | Yes | No | No |
| Pancreatic | Fluorouracil,Gemcitabine,Irinotecan,Oxaliplatin | No | Yes | No | No |
| Pancreatic | Fluorouracil,Gemcitabine,Irinotecan,Oxaliplatin,Paclitaxel Protein-Bound | No | Yes | No | No |
| Pancreatic | Fluorouracil,Gemcitabine,Irinotecan,Paclitaxel Protein-Bound | No | Yes | No | No |
| Pancreatic | Fluorouracil,Gemcitabine,Leucovorin | No | Yes | No | No |
| Pancreatic | Fluorouracil,Gemcitabine,Leucovorin,Oxaliplatin | No | Yes | No | No |
| Pancreatic | Fluorouracil,Gemcitabine,Leucovorin,Oxaliplatin,Paclitaxel Protein-Bound | No | Yes | No | No |
| Pancreatic | Fluorouracil,Gemcitabine,Oxaliplatin,Paclitaxel Protein-Bound | No | Yes | No | No |
| Pancreatic | Fluorouracil,Gemcitabine,Paclitaxel Protein-Bound | No | Yes | No | No |
| Pancreatic | Fluorouracil,Irinotecan Liposomal | No | Yes | No | No |
| Pancreatic | Fluorouracil,Irinotecan Liposomal,Leucovorin | No | Yes | No | No |
| Pancreatic | Fluorouracil,Irinotecan Liposomal,Leucovorin,Oxaliplatin | No | Yes | No | No |
| Pancreatic | Fluorouracil,Irinotecan,Irinotecan Liposomal,Leucovorin | No | Yes | No | No |
| Pancreatic | Fluorouracil,Irinotecan,Leucovorin,Leuprolide,Oxaliplatin | No | Yes | No | No |
| Pancreatic | Fluorouracil,Irinotecan,Leucovorin,Methotrexate,Oxaliplatin | No | Yes | No | No |
| Pancreatic | Fluorouracil,Irinotecan,Leucovorin,Olaparib,Oxaliplatin,Pembrolizumab | No | Yes | No | No |
| Pancreatic | Fluorouracil,Irinotecan,Leucovorin,Oxaliplatin,Paclitaxel Protein-Bound | No | Yes | No | No |
| Pancreatic | Fluorouracil,Irinotecan,Leucovorin,Oxaliplatin,Pembrolizumab | No | Yes | No | No |
| Pancreatic | Fluorouracil,Irinotecan,Leucovorin,Oxaliplatin,Rituximab | No | Yes | No | No |
| Pancreatic | Fluorouracil,Irinotecan,Leucovorin,Oxaliplatin,Trastuzumab | No | Yes | No | No |
| Pancreatic | Fluorouracil,Irinotecan,Leucovorin,Trastuzumab | No | Yes | No | No |
| Pancreatic | Fluorouracil,Irinotecan,Olaparib,Oxaliplatin,Rucaparib | No | Yes | No | No |
| Pancreatic | Fluorouracil,Irinotecan,Oxaliplatin,Ramucirumab | No | Yes | No | No |
| Pancreatic | Fluorouracil,Leucovorin | No | Yes | No | No |
| Pancreatic | Fluorouracil,Leucovorin,Mitomycin | No | Yes | No | No |
| Pancreatic | Fluorouracil,Leucovorin,Nintedanib,Oxaliplatin | No | Yes | No | No |
| Pancreatic | Fluorouracil,Leucovorin,Olaparib,Oxaliplatin | No | Yes | No | No |
| Pancreatic | Fluorouracil,Leucovorin,Oxaliplatin,Paclitaxel Protein-Bound | No | Yes | No | No |
| Pancreatic | Fluorouracil,Leucovorin,Oxaliplatin,Pembrolizumab | No | Yes | No | No |
| Pancreatic | Fluorouracil,Paclitaxel Protein-Bound | No | Yes | No | No |
| Pancreatic | FOLFIRI | No | Yes | No | No |
| Pancreatic | FOLFIRINOX | No | Yes | No | No |
| Pancreatic | FOLFOX | No | Yes | No | No |
| Pancreatic | Fulvestrant,Gemcitabine,Paclitaxel Protein-Bound | No | Yes | No | No |
| Pancreatic | Gemcitabine | No | Yes | No | No |
| Pancreatic | Gemcitabine,Hydroxyurea,Paclitaxel Protein-Bound | No | Yes | No | No |
| Pancreatic | Gemcitabine,Imatinib,Paclitaxel Protein-Bound | No | Yes | No | No |
| Pancreatic | Gemcitabine,Interferon Alfa-2B,Paclitaxel Protein-Bound | No | Yes | No | No |
| Pancreatic | Gemcitabine,Irinotecan | No | Yes | No | No |
| Pancreatic | Gemcitabine,Letrozole,Paclitaxel Protein-Bound | No | Yes | No | No |
| Pancreatic | Gemcitabine,Leucovorin,Paclitaxel Protein-Bound | No | Yes | No | No |
| Pancreatic | Gemcitabine,Nivolumab,Paclitaxel Protein-Bound | No | Yes | No | No |
| Pancreatic | Gemcitabine,Oxaliplatin | No | Yes | No | No |
| Pancreatic | Gemcitabine,Oxaliplatin,Paclitaxel Protein-Bound | No | Yes | No | No |
| Pancreatic | Gemcitabine,Oxaliplatin,Rituximab/Hyaluronidase | No | Yes | No | No |
| Pancreatic | Gemcitabine,Paclitaxel | No | Yes | No | No |
| Pancreatic | Gemcitabine,Paclitaxel Protein-Bound | No | Yes | No | No |
| Pancreatic | Gemcitabine,Paclitaxel Protein-Bound,Pembrolizumab | No | Yes | No | No |
| Pancreatic | Gemcitabine,Pembrolizumab | No | Yes | No | No |
| Pancreatic | Gemcitabine,Trametinib | No | Yes | No | No |
| Pancreatic | Ibrutinib | No | No |  |  |
| Pancreatic | Ipilimumab,Nivolumab | No | No |  |  |
| Pancreatic | Irinotecan | No | Yes | No | No |
| Pancreatic | Irinotecan Liposomal | No | Yes | No | No |
| Pancreatic | Irinotecan,Leucovorin,Oxaliplatin | No | Yes | No | No |
| Pancreatic | Irinotecan,Oxaliplatin | No | Yes | No | No |
| Pancreatic | Lenalidomide | No | No |  |  |
| Pancreatic | Letrozole | No | No |  |  |
| Pancreatic | Leucovorin | No | No |  |  |
| Pancreatic | Leucovorin,Oxaliplatin | No | Yes | No | No |
| Pancreatic | Leuprolide | No | No |  |  |
| Pancreatic | Methotrexate | No | No |  |  |
| Pancreatic | Mitomycin | No | No |  |  |
| Pancreatic | Nivolumab | No | No |  |  |
| Pancreatic | Olaparib | No | Yes | No | No |
| Pancreatic | Oxaliplatin | No | Yes | No | No |
| Pancreatic | Paclitaxel | No | No |  |  |
| Pancreatic | Paclitaxel Protein-Bound | No | Yes | No | No |
| Pancreatic | Paclitaxel Protein-Bound,Vinorelbine | No | Yes | No | No |
| Pancreatic | Palbociclib | No | No |  |  |
| Pancreatic | Pembrolizumab | No | Yes | No | No |
| Pancreatic | Pertuzumab,Trastuzumab | No | No |  |  |
| Pancreatic | Regorafenib | No | No |  |  |
| Pancreatic | Rituximab | No | No |  |  |
| Pancreatic | Rucaparib | No | No |  |  |
| Pancreatic | Ruxolitinib | No | No |  |  |
| Pancreatic | Sorafenib | No | No |  |  |
| Pancreatic | Sunitinib | No | No |  |  |
| Pancreatic | Tamoxifen | No | No |  |  |
| Pancreatic | Trametinib | No | No |  |  |
| Pancreatic | Venetoclax | No | No |  |  |

### eTable 3. Population characteristics for Treatment Selection Analysis

|  | **2019** | | | | |  | **2020** | | | | |  | **Total** | |
| --- | --- | --- | --- | --- | --- | --- | --- | --- | --- | --- | --- | --- | --- | --- |
|  | Jan 1-Mar 8 | | Apr 8-Jul 31 | | SMD |  | Jan 1-Mar 8 | | Apr 8-Jul 31 | | SMD |  |  |  |
| **n** | 1246 |  | 1949 |  |  |  | 1121 |  | 1646 |  |  |  | 5962 |  |
| **Cancer (n, %)** |  |  |  |  |  |  |  |  |  |  |  |  |  |  |
| Breast | 238 | (19.1%) | 391 | (20.1%) | 0.079 |  | 194 | (17.3%) | 278 | (16.9%) | 0.070 |  | 1101 | (18.5%) |
| NSCLC | 715 | (57.4%) | 1146 | (58.8%) |  |  | 684 | (61.0%) | 973 | (59.1%) |  |  | 3518 | (59.0%) |
| Prostate | 191 | (15.3%) | 246 | (12.6%) |  |  | 140 | (12.5%) | 245 | (14.9%) |  |  | 822 | (13.8%) |
| UCC | 102 | (8.2%) | 166 | (8.5%) |  |  | 103 | (9.2%) | 150 | (9.1%) |  |  | 521 | (8.7%) |
| **Median Age (IQR)** | 70 | (63, 77) | 71 | (62, 78) | 0.023 |  | 69 | (62, 77) | 70 | (63, 78) | 0.073 |  | 70 | (63, 78) |
| **Gender (n, %)** |  |  |  |  |  |  |  |  |  |  |  |  |  |  |
| Female | 557 | (44.7%) | 964 | (49.5%) | 0.095 |  | 536 | (47.8%) | 768 | (46.7%) | 0.023 |  | 2825 | (47.4%) |
| Male | 689 | (55.3%) | 985 | (50.5%) |  |  | 585 | (52.2%) | 878 | (53.3%) |  |  | 3137 | (52.6%) |
| **Race (n, %)** |  |  |  |  |  |  |  |  |  |  |  |  |  |  |
| Non-hispanic White | 801 | (71.8%) | 1246 | (71.9%) | 0.014 |  | 679 | (69.1%) | 1016 | (69.7%) | 0.059 |  | 3742 | (70.8%) |
| Non-Hispanic Black | 109 | (9.8%) | 170 | (9.8%) |  |  | 112 | (11.4%) | 141 | (9.7%) |  |  | 532 | (10.1%) |
| Hispanic | 58 | (5.2%) | 85 | (4.9%) |  |  | 45 | (4.6%) | 70 | (4.8%) |  |  | 258 | (4.9%) |
| Other | 147 | (13.2%) | 231 | (13.3%) |  |  | 146 | (14.9%) | 230 | (15.8%) |  |  | 754 | (14.3%) |
| Missing | 131 |  | 217 |  |  |  | 139 |  | 189 |  |  |  | 676 |  |
| **Insurance (n, %)** |  |  |  |  |  |  |  |  |  |  |  |  |  |  |
| Commercial | 597 | (47.9%) | 973 | (49.9%) | 0.07 |  | 575 | (51.3%) | 817 | (49.6%) | 0.044 |  | 2962 | (49.7%) |
| Government | 263 | (21.1%) | 434 | (22.3%) |  |  | 243 | (21.7%) | 352 | (21.4%) |  |  | 1292 | (21.7%) |
| Unknown/not documented/self-pay | 386 | (31.0%) | 542 | (27.8%) |  |  | 303 | (27.0%) | 477 | (29.0%) |  |  | 1708 | (28.6%) |
| **Practice type (n, %)** |  |  |  |  |  |  |  |  |  |  |  |  |  |  |
| Academic | 95 | (7.6%) | 154 | (7.9%) | 0.01 |  | 66 | (5.9%) | 104 | (6.3%) | 0.018 |  | 419 | (7.0%) |
| Community | 1151 | (92.4%) | 1795 | (92.1%) |  |  | 1055 | (94.1%) | 1542 | (93.7%) |  |  | 5543 | (93.0%) |
| **ECOG performance status (n, %)** |  |  |  |  |  |  |  |  |  |  |  |  |  |  |
| 0-1 | 543 | (83.5%) | 780 | (81.4%) | 0.056 |  | 505 | (82.9%) | 728 | (82.0%) | 0.025 |  | 2556 | (82.3%) |
| >=2 | 107 | (16.5%) | 178 | (18.6%) |  |  | 104 | (17.1%) | 160 | (18.0%) |  |  | 549 | (17.7%) |
| Missing | 596 |  | 991 | (50.8%) |  |  | 512 | (45.7%) | 758 | (46.1%) |  |  | 2857 | (47.9%) |
| **Opioid prescription (n, %)** | 77 | (6.2%) | 120 | (6.2%) | 0.001 |  | 63 | (5.6%) | 90 | (5.5%) | 0.007 |  | 350 | (5.9%) |
| **De novo metastatic (n, %)** | 638 | (57.3%) | 1057 | (60.5%) | 0.066 |  | 613 | (60.2%) | 967 | (65.6%) | 0.113 |  | 3275 | (61.2%) |
| Missing | 132 |  | 202 |  |  |  | 102 |  | 172 |  |  |  | 608 |  |

^a^ NSCLC = Non-small cell lung carcinoma; UCC = urothelial cell carcinoma; ECOG = Eastern Cooperative Oncology Group

### eTable 4. Adjusted probabilities of treatment within 30 days by race/ethnicity

|  | **2019** | | | | | | | | | | |  | **2020** | | | | | | | | | | |  | **Difference in differences** | | |
| --- | --- | --- | --- | --- | --- | --- | --- | --- | --- | --- | --- | --- | --- | --- | --- | --- | --- | --- | --- | --- | --- | --- | --- | --- | --- | --- | --- |
|  | Jan 1-Mar 8 | | |  | Apr 8-Jul 31 | | |  | 2019 Diff | | |  | Jan 1-Mar 8 | | |  | Apr 8-Jul 31 | | |  | Diff | | |  | 2020-2019 | | |
|  | Est | (95% CI) | |  | Est | (95% CI) | |  | Est | (95% CI) | |  | Est | (95% CI) | |  | Est | (95% CI) | |  | Est | (95% CI) | |  | Est | (95% CI) | |
| Non-hispanic White | 0.422 | (0.308, | 0.536) |  | 0.429 | (0.318, | 0.540) |  | 0.007 | (-0.027, | 0.041) |  | 0.446 | (0.282, | 0.610) |  | 0.468 | (0.359, | 0.577) |  | 0.022 | (-0.056, | 0.099) |  | 0.015 | (-0.062, | 0.092) |
| Non-Hispanic Black | 0.395 | (0.103, | 0.687) |  | 0.386 | (0.204, | 0.568) |  | -0.009 | (-0.150, | 0.133) |  | 0.460 | (0.275, | 0.646) |  | 0.431 | (0.173, | 0.690) |  | -0.029 | (-0.166, | 0.108) |  | -0.021 | (-0.285, | 0.244) |
| Hispanic | 0.405 | (0.215, | 0.596) |  | 0.414 | (0.286, | 0.542) |  | 0.009 | (-0.105, | 0.122) |  | 0.395 | (0.209, | 0.581) |  | 0.466 | (0.206, | 0.727) |  | 0.071 | (-0.038, | 0.180) |  | 0.063 | (-0.122, | 0.247) |
| Other | 0.411 | (0.177, | 0.645) |  | 0.440 | (0.259, | 0.622) |  | 0.029 | (-0.057, | 0.116) |  | 0.450 | (0.162, | 0.738) |  | 0.490 | (0.265, | 0.714) |  | 0.039 | (-0.045, | 0.124) |  | 0.010 | (-0.050, | 0.070) |

^a^ Results are from race:period interaction model.

### eTable 5. Adjusted probabilities of treatment within 30 days by age

|  | **2019** | | | | | | | | | | |  | **2020** | | | | | | | | | | |  | **Difference in differences** | | |
| --- | --- | --- | --- | --- | --- | --- | --- | --- | --- | --- | --- | --- | --- | --- | --- | --- | --- | --- | --- | --- | --- | --- | --- | --- | --- | --- | --- |
|  | Jan 1-Mar 8 | | |  | Apr 8-Jul 31 | | |  | 2019 Diff | | |  | Jan 1-Mar 8 | | |  | Apr 8-Jul 31 | | |  | Diff | | |  | 2020-2019 | | |
|  | Est | (95% CI) | |  | Est | (95% CI) | |  | Est | (95% CI) | |  | Est | (95% CI) | |  | Est | (95% CI) | |  | Est | (95% CI) | |  | Est | (95% CI) | |
| Age >75 | 0.379 | (0.279, | 0.479) |  | 0.388 | (0.304, | 0.471) |  | 0.009 | (-0.043, | 0.061) |  | 0.412 | (0.254, | 0.570) |  | 0.445 | (0.321, | 0.570) |  | 0.033 | (-0.028, | 0.094) |  | 0.024 | (-0.042, | 0.091) |
| Age <=75 | 0.432 | (0.308, | 0.556) |  | 0.441 | (0.326, | 0.557) |  | 0.009 | (-0.043, | 0.060) |  | 0.459 | (0.309, | 0.609) |  | 0.478 | (0.339, | 0.616) |  | 0.019 | (-0.025, | 0.062) |  | 0.010 | (-0.043, | 0.062) |

^a^Results are from age:period interaction model

### eTable 6. Adjusted probabilities of receiving myelosuppressive therapy by race/ethnicity

|  | **2019** | | | | | | | | | | |  | **2020** | | | | | | | | | | |  | **Difference in differences** | | |
| --- | --- | --- | --- | --- | --- | --- | --- | --- | --- | --- | --- | --- | --- | --- | --- | --- | --- | --- | --- | --- | --- | --- | --- | --- | --- | --- | --- |
|  | Jan 1-Mar 8 | | |  | Apr 8-Jul 31 | | |  | 2019 Diff | | |  | Jan 1-Mar 8 | | |  | Apr 8-Jul 31 | | |  | Diff | | |  | 2020-2019 | | |
|  | Est | (95% CI) | |  | Est | (95% CI) | |  | Est | (95% CI) | |  | Est | (95% CI) | |  | Est | (95% CI) | |  | Est | (95% CI) | |  | Est | (95% CI) | |
| Non-hispanic White | 0.705 | (0.660, | 0.751) |  | 0.672 | (0.620, | 0.724) |  | -0.034 | (-0.072, | 0.005) |  | 0.684 | (0.653, | 0.714) |  | 0.683 | (0.651, | 0.716) |  | 0.000 | (-0.031, | 0.030) |  | 0.033 | (-0.017, | 0.084) |
| Non-Hispanic Black | 0.711 | (0.639, | 0.783) |  | 0.696 | (0.613, | 0.778) |  | -0.016 | (-0.102, | 0.071) |  | 0.706 | (0.639, | 0.773) |  | 0.660 | (0.584, | 0.735) |  | -0.047 | (-0.118, | 0.025) |  | -0.031 | (-0.134, | 0.072) |
| Hispanic | 0.696 | (0.591, | 0.801) |  | 0.711 | (0.625, | 0.798) |  | 0.015 | (-0.094, | 0.125) |  | 0.662 | (0.559, | 0.764) |  | 0.586 | (0.491, | 0.682) |  | -0.075 | (-0.193, | 0.042) |  | -0.091 | (-0.245, | 0.063) |
| Other | 0.657 | (0.566, | 0.748) |  | 0.608 | (0.461, | 0.755) |  | -0.049 | (-0.157, | 0.059) |  | 0.667 | (0.576, | 0.758) |  | 0.629 | (0.524, | 0.734) |  | -0.038 | (-0.092, | 0.016) |  | 0.011 | (-0.104, | 0.126) |

^a^ Results are from race:period interaction model.

### eTable 7. Adjusted probabilities of receiving myelosuppressive therapy by age

|  | **2019** | | | | | | | | | | |  | **2020** | | | | | | | | | | |  | **Difference in differences** | | |
| --- | --- | --- | --- | --- | --- | --- | --- | --- | --- | --- | --- | --- | --- | --- | --- | --- | --- | --- | --- | --- | --- | --- | --- | --- | --- | --- | --- |
|  | Jan 1-Mar 8 | | |  | Apr 8-Jul 31 | | |  | 2019 Diff | | |  | Jan 1-Mar 8 | | |  | Apr 8-Jul 31 | | |  | Diff | | |  | 2020-2019 | | |
|  | Est | (95% CI) | |  | Est | (95% CI) | |  | Est | (95% CI) | |  | Est | (95% CI) | |  | Est | (95% CI) | |  | Est | (95% CI) | |  | Est | (95% CI) | |
| Age >75 | 0.593 | (0.534, | 0.652) |  | 0.542 | (0.469, | 0.616) |  | -0.051 | (-0.109, | 0.007) |  | 0.582 | (0.539, | 0.625) |  | 0.538 | (0.488, | 0.588) |  | -0.044 | (-0.090, | 0.001) |  | 0.007 | (-0.067, | 0.081) |
| Age <=75 | 0.747 | (0.701, | 0.793) |  | 0.726 | (0.671, | 0.781) |  | -0.021 | (-0.058, | 0.016) |  | 0.730 | (0.697, | 0.762) |  | 0.731 | (0.698, | 0.764) |  | 0.001 | (-0.026, | 0.028) |  | 0.022 | (-0.021, | 0.064) |

^a^ Results are from age:period interaction model.

### eTable 8. Adjusted probability of treatment within 30 days using state-specific COVID-19 period exposure dates

|  | **2019** | | | | | | | | | | |  | **2020** | | | | | | | | | | |  | **Difference in differences** | | |
| --- | --- | --- | --- | --- | --- | --- | --- | --- | --- | --- | --- | --- | --- | --- | --- | --- | --- | --- | --- | --- | --- | --- | --- | --- | --- | --- | --- |
|  | Pre SAHO | | |  | Post SAHO | | |  | 2019 Diff | | |  | Pre SAHO | | |  | Post SAHO | | |  | Diff | | |  | 2020-2019 | | |
|  | Est | (95% CI) | |  | Est | (95% CI) | |  | Est | (95% CI) | |  | Est | (95% CI) | |  | Est | (95% CI) | |  | Est | (95% CI) | |  | Est | (95% CI) | |
| Unadjusted | 0.453 | (0.431, | 0.473) |  | 0.432 | (0.417, | 0.447) |  |  |  |  |  | 0.457 | (0.434, | 0.478) |  | 0.470 | (0.453, | 0.486) |  |  |  |  |  |  |  |  |
| Adjusted |  |  |  |  |  |  |  |  |  |  |  |  |  |  |  |  |  |  |  |  |  |  |  |  |  |  |  |
| Combined | 0.424 | (0.336, | 0.512) |  | 0.443 | (0.340, | 0.546) |  | 0.019 | (-0.030, | 0.068) |  | 0.439 | (0.323, | 0.555) |  | 0.483 | (0.379, | 0.587) |  | 0.044 | (0.014, | 0.074) |  | 0.025 | (-0.042, | 0.092) |
| Breast | 0.552 | (0.399, | 0.705) |  | 0.576 | (0.344, | 0.808) |  | 0.024 | (-0.172, | 0.219) |  | 0.607 | (0.427, | 0.787) |  | 0.650 | (0.542, | 0.758) |  | 0.043 | (-0.101, | 0.187) |  | 0.019 | (-0.266, | 0.304) |
| Colorectal | 0.407 | (0.316, | 0.499) |  | 0.424 | (0.313, | 0.535) |  | 0.017 | (-0.082, | 0.116) |  | 0.422 | (0.269, | 0.575) |  | 0.446 | (0.332, | 0.560) |  | 0.024 | (-0.050, | 0.099) |  | 0.007 | (-0.156, | 0.171) |
| NSCLC | 0.400 | (0.313, | 0.487) |  | 0.403 | (0.316, | 0.489) |  | 0.003 | (-0.052, | 0.058) |  | 0.404 | (0.282, | 0.526) |  | 0.450 | (0.325, | 0.574) |  | 0.046 | (0.020, | 0.071) |  | 0.043 | (-0.018, | 0.103) |
| Pancreatic | 0.549 | (0.321, | 0.777) |  | 0.592 | (0.430, | 0.755) |  | 0.044 | (-0.215, | 0.302) |  | 0.533 | (0.389, | 0.677) |  | 0.559 | (0.373, | 0.744) |  | 0.026 | (-0.215, | 0.266) |  | -0.018 | (-0.342, | 0.307) |
| Prostate | 0.362 | (0.201, | 0.524) |  | 0.388 | (0.168, | 0.608) |  | 0.026 | (-0.284, | 0.336) |  | 0.359 | (0.178, | 0.540) |  | 0.454 | (0.312, | 0.595) |  | 0.094 | (-0.148, | 0.337) |  | 0.069 | (-0.465, | 0.602) |
| RCC | 0.353 | (0.207, | 0.499) |  | 0.403 | (0.202, | 0.604) |  | 0.050 | (-0.158, | 0.257) |  | 0.405 | (0.168, | 0.642) |  | 0.465 | (0.340, | 0.590) |  | 0.060 | (-0.108, | 0.227) |  | 0.010 | (-0.307, | 0.327) |
| UCC | 0.357 | (0.242, | 0.472) |  | 0.418 | (0.260, | 0.576) |  | 0.061 | (-0.088, | 0.210) |  | 0.400 | (0.233, | 0.568) |  | 0.441 | (0.310, | 0.571) |  | 0.040 | (-0.151, | 0.232) |  | -0.021 | (-0.244, | 0.202) |

^a^ Results are from cancer type:period interaction model. Washout period defined using state stay-at-home order dates. SAHO = Stay at home order. NSCLC = Non-small cell lung carcinoma; RCC = renal cell carcinoma; UCC = urothelial cell carcinoma

### eTable 9. Adjusted probability of receiving myelosuppressive treatment using state-specific COVID-19 period exposure dates

|  | **2019** | | | | | | | | | | |  | **2020** | | | | | | | | | | |  | **Difference in differences** | | |
| --- | --- | --- | --- | --- | --- | --- | --- | --- | --- | --- | --- | --- | --- | --- | --- | --- | --- | --- | --- | --- | --- | --- | --- | --- | --- | --- | --- |
|  | Pre SAHO | | |  | Post SAHO | | |  | 2019 Diff | | |  | Pre SAHO | | |  | Post SAHO | | |  | Diff | | |  | 2020-2019 | | |
|  | Est | (95% CI) | |  | Est | (95% CI) | |  | Est | (95% CI) | |  | Est | (95% CI) | |  | Est | (95% CI) | |  | Est | (95% CI) | |  | Est | (95% CI) | |
| Unadjusted | 0.687 | (0.651, | 0.723) |  | 0.676 | (0.622, | 0.729) |  | -0.012 | (-0.059, | 0.036) |  | 0.691 | (0.656, | 0.727) |  | 0.658 | (0.626, | 0.690) |  | -0.033 | -(0.062, | -0.004) |  | -0.022 | (-0.081, | 0.038) |
| Adjusted |  |  |  |  |  |  |  |  |  |  |  |  |  |  |  |  |  |  |  |  |  |  |  |  |  |  |  |
| Combined | 0.698 | (0.651, | 0.744) |  | 0.667 | (0.609, | 0.725) |  | -0.031 | (-0.065, | 0.002) |  | 0.683 | (0.651, | 0.714) |  | 0.668 | (0.633, | 0.702) |  | -0.015 | (-0.038, | 0.008) |  | 0.016 | (-0.026, | 0.058) |
| Breast | 0.815 | (0.746, | 0.883) |  | 0.776 | (0.701, | 0.851) |  | -0.039 | (-0.122, | 0.044) |  | 0.827 | (0.783, | 0.870) |  | 0.807 | (0.767, | 0.848) |  | -0.019 | (-0.073, | 0.034) |  | 0.020 | (-0.077, | 0.116) |
| NSCLC | 0.835 | (0.797, | 0.872) |  | 0.808 | (0.761, | 0.856) |  | -0.026 | (-0.071, | 0.018) |  | 0.807 | (0.773, | 0.841) |  | 0.783 | (0.749, | 0.817) |  | -0.024 | (-0.059, | 0.012) |  | 0.003 | (-0.056, | 0.061) |
| Prostate | 0.046 | (0.000, | 0.101) |  | 0.025 | (0.004, | 0.045) |  | -0.022 | (-0.080, | 0.036) |  | 0.025 | (0.008, | 0.042) |  | 0.041 | (0.012, | 0.071) |  | 0.016 | (-0.015, | 0.047) |  | 0.037 | (-0.025, | 0.100) |
| UCC | 0.558 | (0.452, | 0.664) |  | 0.497 | (0.402, | 0.593) |  | -0.061 | (-0.190, | 0.068) |  | 0.583 | (0.505, | 0.660) |  | 0.589 | (0.518, | 0.661) |  | 0.007 | (-0.081, | 0.095) |  | 0.068 | (-0.086, | 0.221) |

^a^ Results are from cancer type:period interaction model. Washout period defined using state stay-at-home order dates. SAHO = Stay at home order. NSCLC = Non-small cell lung carcinoma; UCC = urothelial cell carcinoma
